# Intersections between pneumonia, lowered oxygen saturation percentage and immune activation mediate depression, anxiety and chronic fatigue syndrome-like symptoms due to COVID-19: a nomothetic network approach

**DOI:** 10.1101/2021.06.12.21258815

**Authors:** Hawraa Kadhem Al-Jassas, Hussein Kadhem Al-Hakeim, Michael Maes

**Affiliations:** Department of Chemistry, College of Science, University of Kufa, Iraq; Department of Pharmaceutical Chemistry, Faculty of Pharmacy, University of Kufa, Iraq; School of Medicine, IMPACT-the Institute for Mental and Physical Health and Clinical Translation, Deakin University, Barwon Health, Geelong, Australia; Department of Psychiatry, Medical University of Plovdiv, Plovdiv, Bulgaria; Department of Psychiatry, Faculty of Medicine, Chulalongkorn University, Bangkok, Thailand

**Keywords:** COVID-19, depression, chronic fatigue syndrome, inflammation, neuro-immune, psychiatry.

## Abstract

**Background:** COVID-19 is associated with neuropsychiatric symptoms including increased depressive, anxiety and chronic fatigue-syndrome (CFS)-like physiosomatic (previously known as psychosomatic) symptoms.

**Aims:** To delineate the associations between affective and CFS-like symptoms in COVID-19 and chest CT-scan anomalies (CCTAs), oxygen saturation (SpO_2_), interleukin (IL)-6, IL-10, C-Reactive Protein (CRP), albumin, calcium, magnesium, soluble angiotensin converting enzyme (ACE2) and soluble advanced glycation products (sRAGEs).

**Method:** The above biomarkers were assessed in 60 COVID-19 patients and 30 heathy controls who had measurements of the Hamilton Depression (HDRS) and Anxiety (HAM-A) and the Fibromyalgia and Chronic Fatigue (FF) Rating Scales.

**Results:** Partial Least Squares-SEM analysis showed that reliable latent vectors could be extracted from a) key depressive and anxiety and physiosomatic symptoms (the physio-affective or PA-core), b) IL-6, IL-10, CRP, albumin, calcium, and sRAGEs (the immune response core); and c) different CCTAs (including ground glass opacities, consolidation, and crazy paving) and lowered SpO2% (lung lesions). PLS showed that 70.0% of the variance in the PA-core was explained by the regression on the immune response and lung lesions latent vectors. Moreover, one common “infection-immune-inflammatory (III) core” underpins pneumonia-associated CCTAs, lowered SpO2 and immune activation, and this III core explains 70% of the variance in the PA core, and a relevant part of the variance in melancholia, insomnia, and neurocognitive symptoms.

**Discussion:** Acute SARS-CoV-2 infection is accompanied by lung lesions and lowered SpO2 which both may cause activated immune-inflammatory pathways, which mediate the effects of the former on the PA-core and other neuropsychiatric symptoms due to SARS-CoV-2 infection.

## Introduction

SARS coronavirus 2 (SARS-CoV-2) affected more than 157 million people worldwide as of late November 2020, with more than 3.27 million deaths until May, 2021 (Coronavirus-Resource-Center, 2021). SARS-CoV-2 infection has a broad clinical scope, ranging from asymptomatic infection, mild sickness, moderate upper respiratory tract disease, to severe viral pneumonia with respiratory failure and even death (Krishnan et al., 2021; Montenegro et al., 2021; Zhou et al., 2020). Chest imaging, especially computed tomography scan (CT-scan), is critical for the diagnosis, management, and follow-up of COVID-19 infection (Fang Y, 2020; Zhang et al., 2020a). CT scan anomalies (CCTAs), including ground-glass opacities (GGOs), pulmonary densification areas consistent with residual lesions, pneumonic consolidation, and crazy-paving trends are observed in 78.3% of RT-PCR test–proven COVID-19 patients and are associated with lower peripheral oxygen saturation (SpO2) (Al-Hakeim et al., 2021).

SARS-CoV-2 may cause an exaggerated host immune response which may result in lung pathology (Huang et al., 2020; Hui and Zumla, 2019) and organ dysfunctions which are caused by direct virus-induced tissue injury, a systemic inflammatory response and the synergistic effects of both (Darif et al., 2021). The severe lung injury and pulmonary inflammation due to COVID-19 are accompanied by elevated levels of pro-inflammatory cytokines including interleukin-6 (IL-6) and chemokines (Liu et al., 2020a; Mehta et al., 2020). IL-6 is one of the cytokines which causes the acute phase response with increased levels of positive and negative acute phase proteins (including increased levels of C-reactive protein C (CRP) and lowered levels of albumin (Tanaka et al., 2014). SARS-CoV- 2 also causes the release of T-helper (Th)-1 pro-inflammatory and Th-2 anti-inflammatory an T- regulatory cytokines, such as IL-10, which has protective properties against lung injury (Huang et al., 2020) (Lindner et al., 2021). COVID-19 and increased CCTAs are accompanied by increased serum IL-6, CRP, and IL-10, and lowered albumin and oxygen saturation percentage (SpO_2_) (Al-Hakeim et al., 2021). Moreover, we detected increased levels of sRAGE (soluble receptor for advanced glycation end-products) and angiotensin converting enzyme 2 (ACE2) in COVID-19 patients (Al-Hakeim et al., 2021). sRAGEs are generated through proteolysis of the extracellular domain of RAGE or through alternative RNA splicing (Sterenczak et al., 2009; Zhang et al., 2008). Binding of AGEs to membrane RAGEs initiates the transcription of pro-inflammatory transcription factors (Macaione et al., 2007; Tobon-Velasco et al., 2014) with consequent production of IL-6 and other inflammatory mediators (Wang and Liu, 2016). sACE2 is cleaved from membrane-associated ACE2 and is consequently released into the extracellular environment (Lambert et al., 2005). The COVID-19 virus may bind with high affinity to human cells via ACE2 receptors leading to endocytosis of the virus (Pouya et al., 2020; Vlachakis et al., 2020).

COVID-19 is frequently associated with mental health symptoms. Depression is present in 27% of the admitted COVID-19 patients, anxiety in 67%, and sleep disorders in 63% (Yadav et al., 2021). Another study reported an increased prevalence of depression (29.2%) in patients who experienced COVID-19 infection (Zhang et al., 2020b). Anxiety levels in patients with COVID-19 are associated with severity of the condition and comorbidities (Yadav et al., 2021). The effects of COVID-19 on mood symptoms are often described as being the consequence of psychological effects. Thus, not only people with COVID-19 but also people who had contact with COVID-19 infected individuals show increased levels of depression and anxiety (Cao et al., 2020; Oxley et al., 2020; Wang et al., 2020). In addition, self-isolation during lockdowns is associated with an increased prevalence of depressive and anxiety symptoms and this was explained by feelings of isolation (Gualano et al., 2020; Xiao et al., 2020a, b). Patients with either major depression or bipolar disorder show increased psychological distress in response to these SARS-CoV-2-associated phenomena (Van Rheenen et al., 2020). Xiang et al. (Xiang et al., 2020) reported that COVID-19 patients are more likely to experience neuropsychiatric syndromes because of the stigma associated with the illness and anxiety over the infection’s effect. Kornilaki (2021) (Kornilaki, 2021) reported that increased levels of depression, negative affect and anxiety as a results of the COVID-19 symptoms or due to the quarantine state.

Nevertheless, there is now evidence that mood disorders including depression and anxiety have an organic substrate and are characterized by activated immune-inflammatory pathways (Maes et al., 1990; Maes and Carvalho, 2018) including increased levels of proinflammatory (e.g., IL-6) and anti-inflammatory (e.g., IL-10) cytokines, and an acute phase response as indicated by higher levels of CRP and lowered levels of albumin (Maes, 1993; Maes et al., 1993). There is also evidence that these neuro- immune pathways have detrimental effects on gray and white matter neuroplasticity, thereby inducing the biobehavioral changes characteristic of mood disorders (Leonard and Maes, 2012). Therefore, it is appropriate to posit that mood symptoms due to COVID-19 are mediated at least in part by neuro- immune pathways. People with COVID-19 also frequently suffer from mental fatigue, physical fatigue, mild loss of concentration, neurocognitive deficits, headache and myalgia (Borges do Nascimento et al., 2020; Liu et al., 2020b; Zhang et al., 2020c; Zhu et al., 2020), symptoms which are reminiscent of Myalgic Encephalomyelitis / chronic fatigue syndrome (ME/CFS) (Maes and Twisk, 2010). As with mood disorders, patients with ME/CFS show activated neuro-immune pathways with increased levels of pro- and anti-inflammatory cytokines, an acute phase response and multiple signs of nitro-oxidative damage (Bjørklund et al., 2020b; Morris and Maes, 2013). Nevertheless, no research has delineated the immune pathways of affective and ME/CFS-like symptoms in people with COVID-19.

Hence, the aim of the present study was to delineate the associations between affective and ME/CFS-like symptoms and a) CCTAs and SpO_2_, and b) IL-6, IL-10, CRP, albumin, calcium, magnesium, sACE2, and sRAGE in COVID-19. The specific hypothesis is that mood and ME/CFS- like symptoms in COVID-19 are significantly and positively associated with CCTAs, IL-6, IL-10, CRP, sACE2 and sRAGEs, and negatively with SpO_2_, albumin, calcium, and magnesium.

### Subjects and Methods

#### Subjects

Between September and November 2020, sixty COVID-19 male patients aged 25 to 59 years were recruited at the Al-Sadr Teaching Hospital and Al-Amal Specialized Hospital for Communicable Diseases in Najaf governorate, Iraq. These hospitals are official quarantine facilities specializing in COVID-19 care in Iraq. All patients had acute respiratory syndrome (ARS) and were diagnosed with SARS-CoV-2 infection based on positive COVID-19 nucleic acid findings by reverse transcription real-time polymerase chain reaction (rRT-PCR), positive IgM, and ARS disease symptoms including fever, breathing problems, cough, anosmia, and ageusia. The normal controls were males matched for age with the patients. We excluded controls and patients with pre-existing medical conditions, such as type 1 diabetes, and liver, kidney, and cardiovascular disease and cancer, and pre-existing neuro- psychiatric disease including dementia, Parkinson’s disorder, multiple sclerosis, and axis-I psychiatric disorders including major depressive disorder, bipolar disorder, generalized anxiety disorder, and schizophrenia. The study was approved by the institutional ethics board of the University of Kufa (617/2020). Before taking part in this study, all participants and the guardians of COVID-19 patients gave written informed consent. The work was carried out in compliance with Iraqi and foreign ethics and privacy rules, as well as the World Medical Association Declaration of Helsinki, The Belmont Report, CIOMS Guideline, and International Conference on Harmonization of Good Clinical Practice, our IRB adheres to the International Guideline for Human Research Safety (ICH-GCP).

#### Clinical Measurements

A senior psychiatrist assessed the 21-item Hamilton Depression Rating Scale (HDRS) score (Hamilton, 1960). We assessed the first 17 items to measure of severity of illness, while item 18 (diurnal variation) was used in composite scores (see below). Severity of anxiety symptoms was measured employing the Hamilton Anxiety Rating Scale (HAM-A) (Hamilton, 1959). In addition, three HDRS and two HAM-A subdomain scores were computed as explained previously (Almulla et al., 2021). The HDRS subdomain were a) the key depressive symptom domain (key HDRS), namely the sum of depressed mood + feelings of guilt + suicidal ideation (but without loss of work and activities); b) the physiosomatic symptom domain (physiosomatic HDRS), namely the sum of anxiety somatic + somatic symptoms, gastrointestinal + somatic symptoms, general + genital symptoms + hypochondriasis; and c) the melancholic symptom domain (melancholia HDRS), namely the sum of insomnia late + psychomotor retardation + diurnal variation + loss of weight. The HAM-A subdomain scores were a) the key anxiety symptom domain (key HAM-A), namely the sum of anxious mood + tension + fears + anxious behavior at interview; and b) the HAM-A physiosomatic symptom domain (physiosomatic HAM-A), namely the sum of somatic muscular + somatic sensory + cardiovascular symptoms + gastrointestinal symptoms + genitourinary symptoms + autonomic symptoms (but not the respiratory symptoms).

The same senior psychiatrist also assessed the Fibromyalgia and Chronic Fatigue Syndrome Rating (FF) scale (Zachrisson et al., 2002). This scale assesses 12 FF symptoms, namely FF1: muscle pain, FF2: muscular tension, FF3: fatigue, FF4: concentration difficulties, FF5: failing memory, FF6: irritability, FF7: sadness, FF8: sleep disturbances, FF9: autonomic disturbances, FF10: irritable bowel, FF11: headache, and FF12: a flu-like malaise. We used the total sum of all items as an index of overall severity of physio-somatic symptoms (Kanchanatawan et al., 2018). We also computed a purer physiosomatic FF score (physiosomatic FF) as the sum of FF1 + FF2 + FF3 + FF9 + FF10 + FF11 + FF12. Consequently, we computed the sum of all physiosomatic scores, namely z score of physiosomatic HDRS + z physiosomatic HAM-A + z physiosomatic FF. Moreover, we computed z composite scores reflecting cognitive impairments as: z HAM-A item 5 + z FF4 + z FF5. Finally, we also computed an insomnia z composite score as z HDRS items 4, 5 and 6 + z HAMA item 4 + z FF8. The diagnosis of tobacco use disorder (TUD) was made using the DSM-IV-RT criteria. Body mass index (BMI) was determined by dividing weight in kilograms by height in meters squared.

#### Measurements of biomarkers

RT-PCR tests were conducted employing the Applied Biosystems^®^ QuantStudio™ 5 Real- Time PCR System (Thermo Fisher Scientific) supplied by Life Technologies Holdings Pte Ltd. (Marsiling Industrial Estate, Singapore) using the Lyra^®^ Direct SARS-CoV-2 real-time RT-PCR Assay kits (Quidel Corporation, CA, USA). This kit offers a real-time RT-PCR assay for detecting human SARS-CoV-2 in viral RNA isolated from nasal, nasopharyngeal, or oropharyngeal swab specimens. The Assay is designed to detect the SARS-CoV-2 virus’s non-structural Polyprotein (pp1ab). The procedures were carried out as stated in the kit’s instruction manual.

Chest computed tomography anomalies (CCTAs) were measured using the SOMATOM Concept AS (Siemens, Munchen, Germany). We used the world standard nomenclature (Franquet, 2011; Hansell et al., 2008) to assess GGOs, regions of pulmonary densification associated with latent lesions, pneumonic consolidation, and crazy-paving trends (Kwee and Kwee, 2020)

After an overnight fast (at least 10 hours) and before having breakfast, we sampled blood between 7.30 and 9.00 a.m. Venous blood samples (5 mL) were taken and placed in sterile plain tubes. Samples that had been hemolyzed were rejected. The clotted blood samples were centrifuged for five minutes at 3000 rpm after ten minutes, and the serum was removed and transferred to three fresh Eppendorf tubes. IgG and IgM were measured in the sera of patients and controls using a qualitative ACON® COVID-19 IgG/IgM rapid screen. The kits have a 99.1% sensitivity and a 98.2% reliability. We only included patients with positive IgM tests. CRP was measured qualitatively and semi-quantitatively in human serum using a C-Reactive Protein (CRP) latex slide test (Spinreact®, Barcelona, Spain). We used Melsin Medical Co. (Jilin, China) ELISA kits to measure serum IL-6, IL- 10, sRAGE, and sACE2. All analytes showed an inter-assay CV of < 12%. Biolabo^®^, Maizy, France, provided spectrophotometric kits to test total calcium, albumin, and magnesium.

### Statistical analysis

We used analysis of variance (ANOVA) to check whether there were differences in scale variables between diagnostic groups. The analysis of contingency tables (the χ^2^-test) was employed to check whether there were significant associations between nominal variables. To examine the associations between biomarkers and the clinical scores we used correlation matrices based on Pearson’s product-moment correlation coefficients. To delineate the associations between diagnosis and biomarkers, we used univariate generalized linear model (GLM). Consequently, we conducted protected pairwise comparisons among treatment means. False discovery rate p-correction was used to correct for multiple comparisons (Benjamini and Hochberg, 1995). Multiple regression analysis was used to determine the most important biomarkers, which predict the rating scale scores, while allowing for the effects of demographic data (e.g. age and education). We used an automated stepwise method with a p-to-entry of 0.05 and a p-to-remove of 0.06. We checked R^2^ changes, multivariate normality (Cook’s distance and leverage), homoscedasticity (with the White and modified Breusch-Pagan tests), and multicollinearity (using tolerance and variance inflation factor). All results of regression analyses were bootstrapped (5.000 samples) and the latter results are shown if the results are not concordant. All tests were two-tailed, and significance was set at p=0.05. All statistical analyses were performed using IBM SPSS windows version 25, 2017.

We used Smart Partial Least Squares (SmartPLS)-SEM path analysis (Ringle et al., 2012) to assess the multi-step multiple mediated paths between input variables (biomarkers and CCTAs) and the clinical rating scale scores. The latter was entered as a latent vector extracted from the different total and subdomain scores. Output variables that could not be combined in latent vectors were entered as single indicators. The primary input variables were entered (if possible) as one latent vector comprising CCTAs, GGOs, consolidation or other CCTAS, and SpO_2_ (labelled as COVID-19 pneumonia). The biomarker data were considered to (partially) mediate the effects of COVID-19 pneumonia on the symptom domains. The input biomarker variables were (if possible) combined in one latent vector reflecting immune activation (e.g. IL-6, CRP, IL-10, sRAGE, albumin, calcium) and the other biomarkers were entered as single indicators. The latent vectors were conceptualized as reflective models. We performed complete SmartPLS analysis using 5.000 bootstrap samples only when the inner/outer models complied with specific quality data: a) Confirmatory Tetrad analysis confirms that the latent vectors are not mis-specified as reflective models; b) the overall fit of the pathway model is adequate with SRMR < 0.08; c) the outer model latent vector loadings are > 0.666 at p<0.001; and d) the latent vectors show an accurate construct validity as indicated by an average variance extracted (AVE) > 0.5, Cronbach’s alpha > 0.7, rho_A > 0.8, and composite reliability > 0.7. Consequently, we conducted complete PLS path analysis on 5.000 bootstrap samples and computed path coefficients (with p value), outer model loadings, and specific indirect and total effects. We employed Blindfolding and PLSpredict with 10-fold cross-validation to check the predictive performance of the model (Shmueli et al., 2019). Predicted-Oriented Segmentation analysis, Multi- Group Analysis and Measurement Invariance Assessment were used to assess compositional invariance.

### Results

#### Socio-demographic and clinical data

**Table 1** displays the socio-demographic data in the controls and two COVID-19 patient groups divided into those with normal to moderately reduced SpO_2_ values (≥ 76%) and those with extremely low SpO_2_ values (< 76%). Patients with SpO_2_ <76% are somewhat older than the other groups. No significant differences among these study groups were detected in BMI, education, residency, marital status, employment, and TUD. Patients with SpO_2_ < 76% had higher total CCTAs, GGO, consolidation, crazy-paving and other chest abnormalities than COVID patients with SpO_2_ ≥76. The differences in CCTA, crazy-paving and other patters remained significant after FDR p-correction (at p=0.0133). All COVID-19 patients were treated with vitamin C and D and this frequency was higher than in controls. More patients in the SpO_2_ <76% group were treated with dexamethasone than in the > 76% SpO_2_ patient group. There were no significant differences in the treatment with Famotidine, Azithromycin, Meropenem, Heparin, and Clexane between both COVID-19 groups. All COVID-19 patients were on O_2_ therapy and were receiving daily treatment with paracetamol and bromhexine.

**Table 1.**
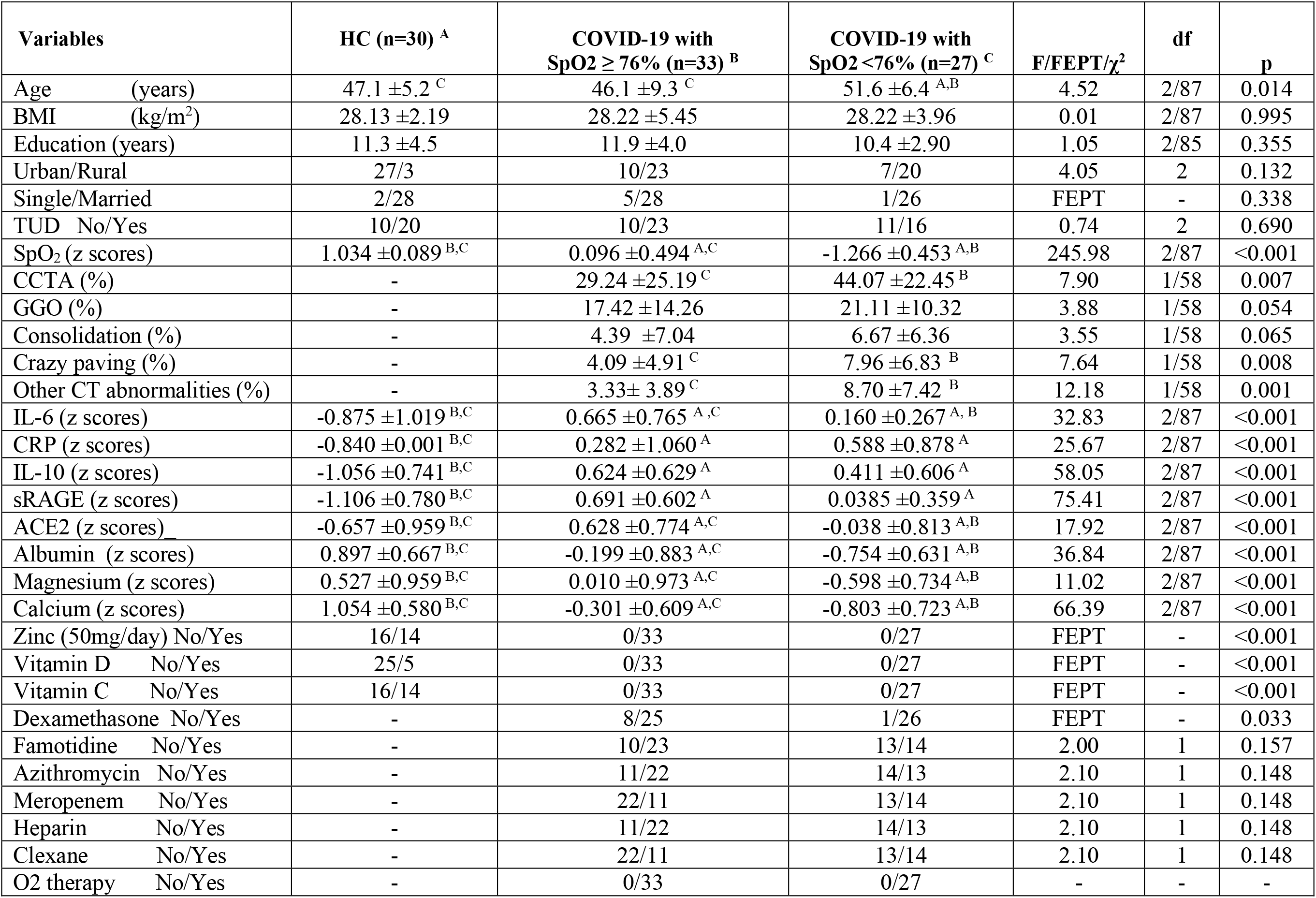
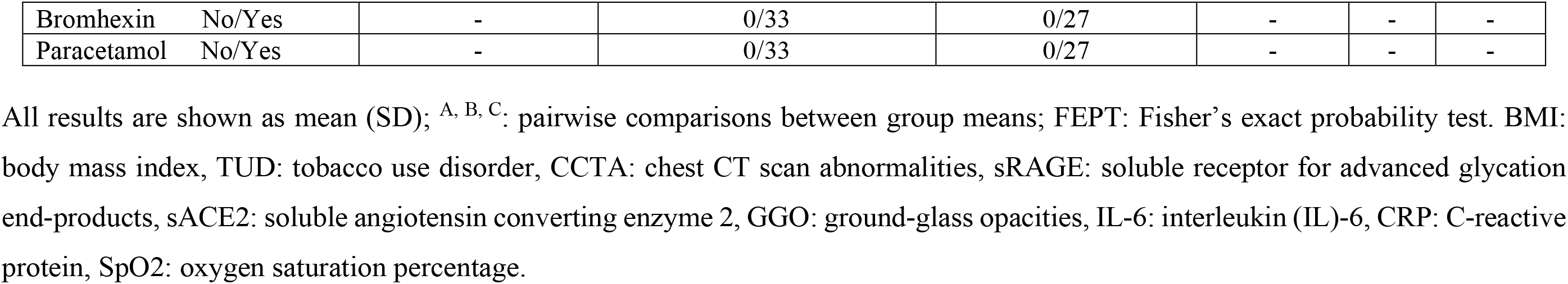
Socio-demographic and clinical data of healthy controls (HC) and COVID-19 patients divided into those with SpO2 values ≥ 76% and with values < 76%.

#### Differences in biomarkers between CIVID-19 subgroup and controls

Table 1 displays the measurements of the various biomarkers in the three study samples and shows substantial differences in all biomarkers, which remained significant after p-correction for FDR (at p=0.0011). There was a significant decrease in serum albumin, calcium, and magnesium in COVID- 19 and increases in IL-6, CRP, IL-10, sRAGE, glucose and ACE2 as compared with normal controls. Moreover, serum IL-6, ACE2, albumin, magnesium, and calcium were significantly reduced in patients with SpO2 <76% as compared with patients with higher SpO_2_ values.

#### Differences in clinical scores among the study groups

**Table 2** shows the measurements of the HDRS, HAM-A and FF total and subdomains scores in both COVID-19 subgroups and controls. All FF and HAM-A total and subdomain scores as well as the sum of all physiosomatic scores, and the cognitive and insomnia scores were significantly higher in COVID-19 patients than in controls. The total HDRS-17 and melancholia HDRS scores were significantly different between the three subgroups and increased from controls → COVID-19 with SpO_2_ ≥ 76%. → COVID-19 with SpO_2_ < 76%.

**Table 2.** Measurements of affective and physiosomatic symptom scores in healthy controls (HC) and COVID-19 patients divided into those with SpO2 values ≥ 76% and with values < 76%.

| Variables | HC<br>(n=30) | COVID-19 with<br>SpO2 $\geq 76\%$ (n=33) <sup>B</sup> | COVID-19 with<br>SpO2 $< 76\%$ (n=27) <sup>C</sup> | F/$\chi^2$ | df | p |
| --- | --- | --- | --- | --- | --- | --- |
| Key HDRS | 0.63 ± 0.67 <sup>B,C</sup> | 2.94 ± 1.44 <sup>A</sup> | 2.74 ± 1.16 <sup>A</sup> | 37.71 | 2/87 | <0.001 |
| Physiosomatic HDRS | 1.83 ± 0.91 <sup>B,C</sup> | 4.52 ± 1.50 <sup>A</sup> | 4.82 ± 1.62 <sup>A</sup> | 42.32 | 2/87 | <0.001 |
| Melancholia HDRS | 0.70 ± 0.84 <sup>B,C</sup> | 2.24 ± 1.39 <sup>A,C</sup> | 2.96 ± 1.43 <sup>A,B</sup> | 24.88 | 2/87 | <0.001 |
| Total HDRS | 5.17 ± 1.56 <sup>B,C</sup> | 15.73 ± 2.97 <sup>A,C</sup> | 17.63 ± 2.17 <sup>A,B</sup> | 243.65 | 2/87 | <0.001 |
| Physiosomatic FF | 2.77 ± 1.14 <sup>B,C</sup> | 13.30 ± 4.10 <sup>A</sup> | 13.59 ± 4.09 <sup>A</sup> | 98.17 | 2/87 | <0.001 |
| Total FF | 6.57 ± 1.61 <sup>B,C</sup> | 22.55 ± 6.48 <sup>A</sup> | 22.85 ± 5.59 <sup>A</sup> | 101.32 | 2/87 | <0.001 |
| Key HAM-A | 0.70 ± 0.75 <sup>B,C</sup> | 4.09 ± 1.49 <sup>A</sup> | 3.70 ± 1.35 <sup>A</sup> | 67.57 | 2/87 | <0.001 |
| Physiosomatic HAM-A | 0.40 ± 0.56 <sup>B,C</sup> | 3.46 ± 1.89 <sup>A</sup> | 3.41 ± 1.62 <sup>A</sup> | 41.72 | 2/87 | <0.001 |
| Total HAM-A | 2.47 ± 1.63 <sup>B,C</sup> | 12.67 ± 3.41 <sup>A</sup> | 12.74 ± 2.61 <sup>A</sup> | 145.55 | 2/87 | <0.001 |
| All physiosomatic (z scores) | -1.24 ± 0.20 <sup>B,C</sup> | 0.59 ± 0.55 <sup>A</sup> | 0.66 ± 0.59 <sup>A</sup> | 151.10 | 2/87 | <0.001 |
| Cognitive symptoms (z scores) | -0.48 ± 0.70 <sup>B,C</sup> | 0.32 ± 1.23 <sup>A</sup> | 0.15 ± 0.76 <sup>A</sup> | 6.04 | 2/87 | 0.003 |
| Insomnia symptoms (z scores) | -0.67 ± 0.70 <sup>B,C</sup> | 0.22 ± 1.10 <sup>A</sup> | 0.48 ± 0.75 <sup>A</sup> | 13.76 | 2/87 | <0.001 |

All results are shown as mean ± SD; <sup>A, B, C</sup>: pairwise comparisons between group means; FF: Fibromyalgia and Chronic Fatigue Syndrome
Rating Scale, HDRS: Hamilton Depression Rating Scale, HAM-A: Hamilton Anxiety Rating Scale.

#### Correlations between rating scale scores and biomarkers

**Table 3** shows the intercorrelation matrix between the total HDRS, FF, and HAM-A scores and CCTAs, SpO_2_, and the biomarkers in the total study group. The HDRS, FF and HAM-A scores showed positive significant associations with CCTAs, CRP, IL-6, IL-10, sRAGE, and ACE2, and inverse correlations with SpO_2_, albumin, magnesium and calcium.

**Table 3.**
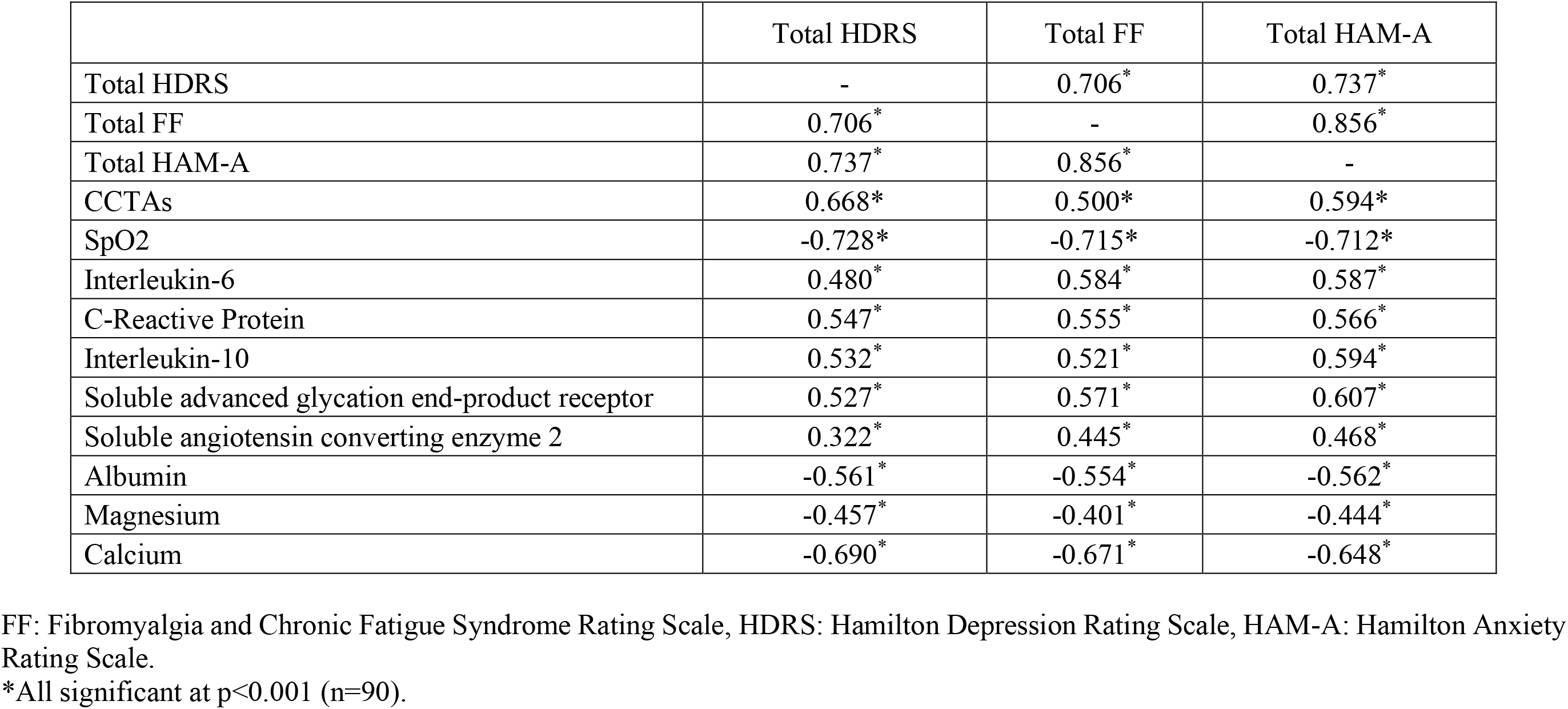
Intercorrelation matrix between affective and physiosomatic rating scale scores, Chest CT scan anomalies (CCTAs), blood oxygen saturation percentage (SpO2), and diverse biomarkers.

#### Prediction of the Hamilton Depression Rating Scale (HDRS) scores

The results of multiple regression of the HDRS total and subdomain scores on the measured biomarkers as dependent variables are presented in **Table 4**. Regression #1 shows that 72.6% of the variance in the total HDRS score could be explained by the regression on sRAGE and CCTA (all positively associated) and SpO_2_ (inversely associated). Regression #2 shows that 65.7% of the variance in the total HDRS score could be explained by regression on sRAGE (positively) and calcium (negatively). A significant part of the variance in the key HDRS scores (39.5%) could be explained by the regression on the IL-6 (positively) and calcium (inversely). Regression #4 shows that 43.3% of the variance in the physiosomatic HDRS symptoms could be explained by the regression on CCTA and IL-10 (both positively). **Figure 1** shows the partial regression plot of the physiosomatic HDRS score on the CCTAs values after adjusting for IL-10. Furthermore, IL-10 and CRP explained 30.8% of the variance in the physiosomatic HDRS score (Regression #5). A significant part of the variance (36.2%) in the melancholia HDRS scores could be explained by the regression on calcium and SpO_2_.

**Figure 1.**
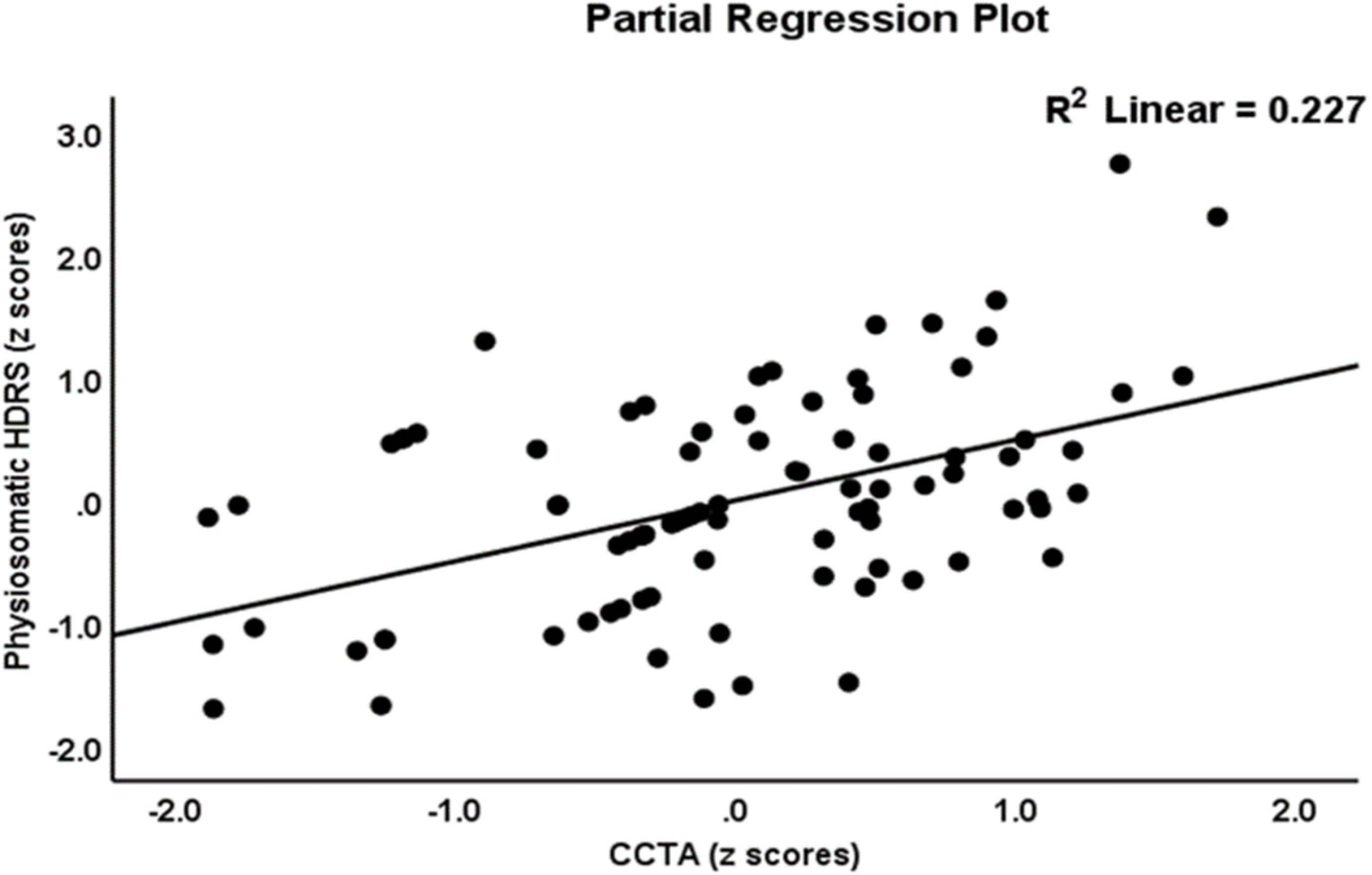
Partial regression of the physiosomatic component of the Hamilton-Depression-Rating-Scale (HDRS) score on the total chest CT scan anomalies (CCTAs).

**Table 4.**
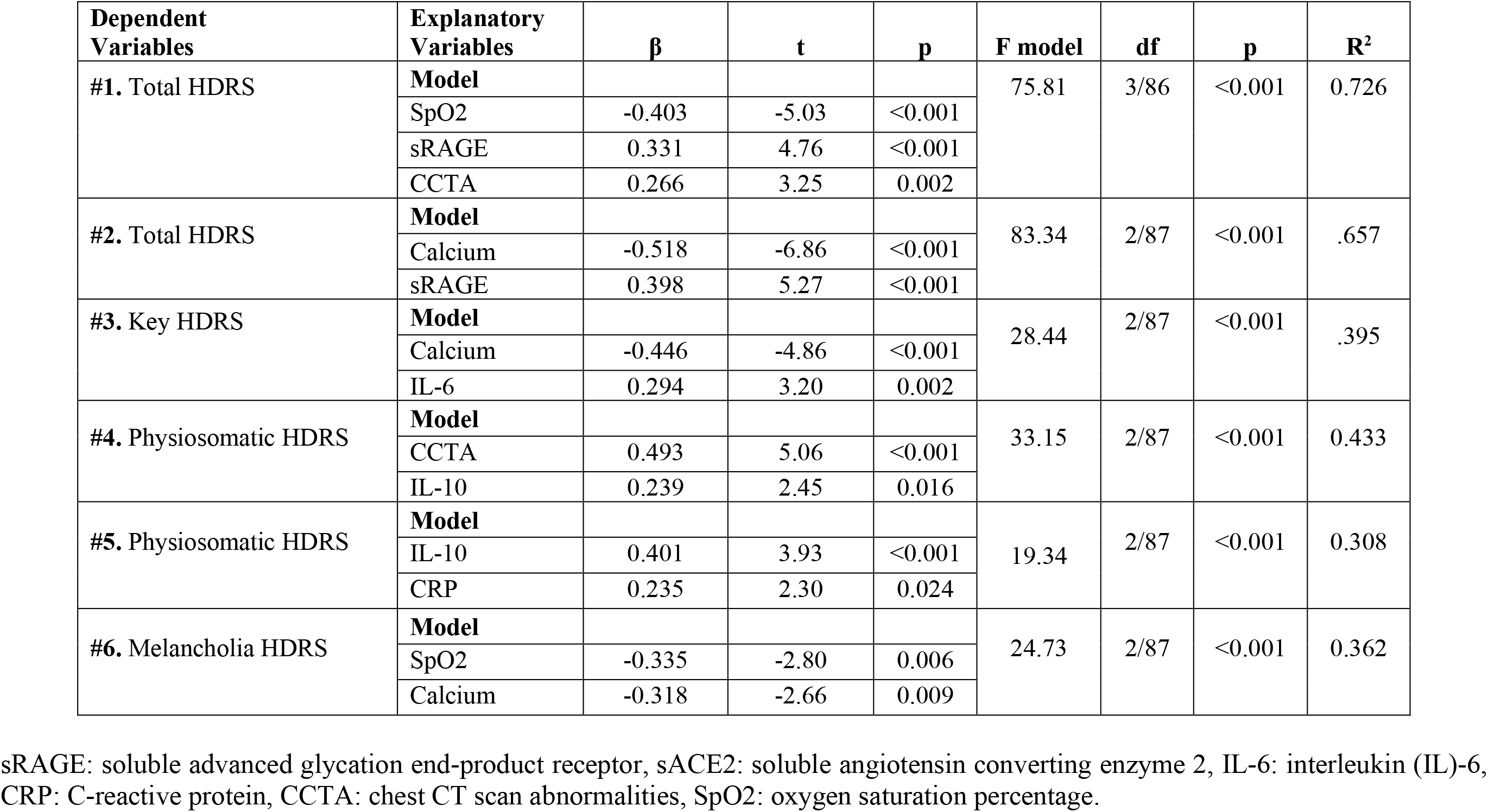
Results of multiple regression analyses with the Hamilton Depression Rating Scale (HDRS), total and subdomain, scores as dependent variables and biomarkers as explanatory variables.

| Dependent Variables | Explanatory Variables | $\beta$ | t | p | F model | df | p | R<sup>2</sup> |
| --- | --- | --- | --- | --- | --- | --- | --- | --- |
| #1. Total HDRS | Model |  |  |  | 75.81 | 3/86 | <0.001 | 0.726 |
|  | SpO2 | -0.403 | -5.03 | <0.001 |  |  |  |  |
|  | sRAGE | 0.331 | 4.76 | <0.001 |  |  |  |  |
|  | CCTA | 0.266 | 3.25 | 0.002 |  |  |  |  |
| #2. Total HDRS | Model |  |  |  | 83.34 | 2/87 | <0.001 | .657 |
|  | Calcium | -0.518 | -6.86 | <0.001 |  |  |  |  |
|  | sRAGE | 0.398 | 5.27 | <0.001 |  |  |  |  |
| #3. Key HDRS | Model |  |  |  | 28.44 | 2/87 | <0.001 | .395 |
|  | Calcium | -0.446 | -4.86 | <0.001 |  |  |  |  |
|  | IL-6 | 0.294 | 3.20 | 0.002 |  |  |  |  |
| #4. Physiosomatic HDRS | Model |  |  |  | 33.15 | 2/87 | <0.001 | 0.433 |
|  | CCTA | 0.493 | 5.06 | <0.001 |  |  |  |  |
|  | IL-10 | 0.239 | 2.45 | 0.016 |  |  |  |  |
| #5. Physiosomatic HDRS | Model |  |  |  | 19.34 | 2/87 | <0.001 | 0.308 |
|  | IL-10 | 0.401 | 3.93 | <0.001 |  |  |  |  |
|  | CRP | 0.235 | 2.30 | 0.024 |  |  |  |  |
| #6. Melancholia HDRS | Model |  |  |  | 24.73 | 2/87 | <0.001 | 0.362 |
|  | SpO2 | -0.335 | -2.80 | 0.006 |  |  |  |  |
|  | Calcium | -0.318 | -2.66 | 0.009 |  |  |  |  |

sRAGE: soluble advanced glycation end-product receptor, sACE2: soluble angiotensin converting enzyme 2, IL-6: interleukin (IL)-6, CRP: C-reactive protein, CCTA: chest CT scan abnormalities, SpO2: oxygen saturation percentage.

#### Prediction of the FF scale scores

**Table 5** shows the results of multiple regression analyses with the FF total and subdomain scores as dependent variables and the biomarkers as explanatory variables. We found that 64.3% of the variance in the total FF score could be explained by the regression on IL-6 and sRAGE (both positively) and SpO2 (negatively) (Regression #1). Regression #2 shows that 59.8% of the variance in the FF total score could be explained by the regression on the sRAGE, CRP, and IL-6 (all positively) and calcium (inversely). Regression #3 showed that 61.6% of the variance in the physiosomatic FF symptoms could be explained by the regression on IL-6 and sRAGE (both inversely) and SpO_2_ (positively). **Figure 2** shows the partial regression plot of the physiosomatic FF score on SpO_2_ after adjusting for IL-6 and sRAGE. Regression #4 showed that 56.3% of the physiosomatic FF symptoms could be explained by the regression on IL- 6 and sRAGE (positively) and calcium (inversely). Regression #5 showed that 46.2% of the variance in the fatigue score could be explained by the regression on sRAGE (positively) and SpO_2_ (inversely). Regression #6 showed that 47.0% of the variance in the fatigue score could be explained by the regression on CRP and sRAGE (positively) and calcium (negatively).

**Figure 2.**
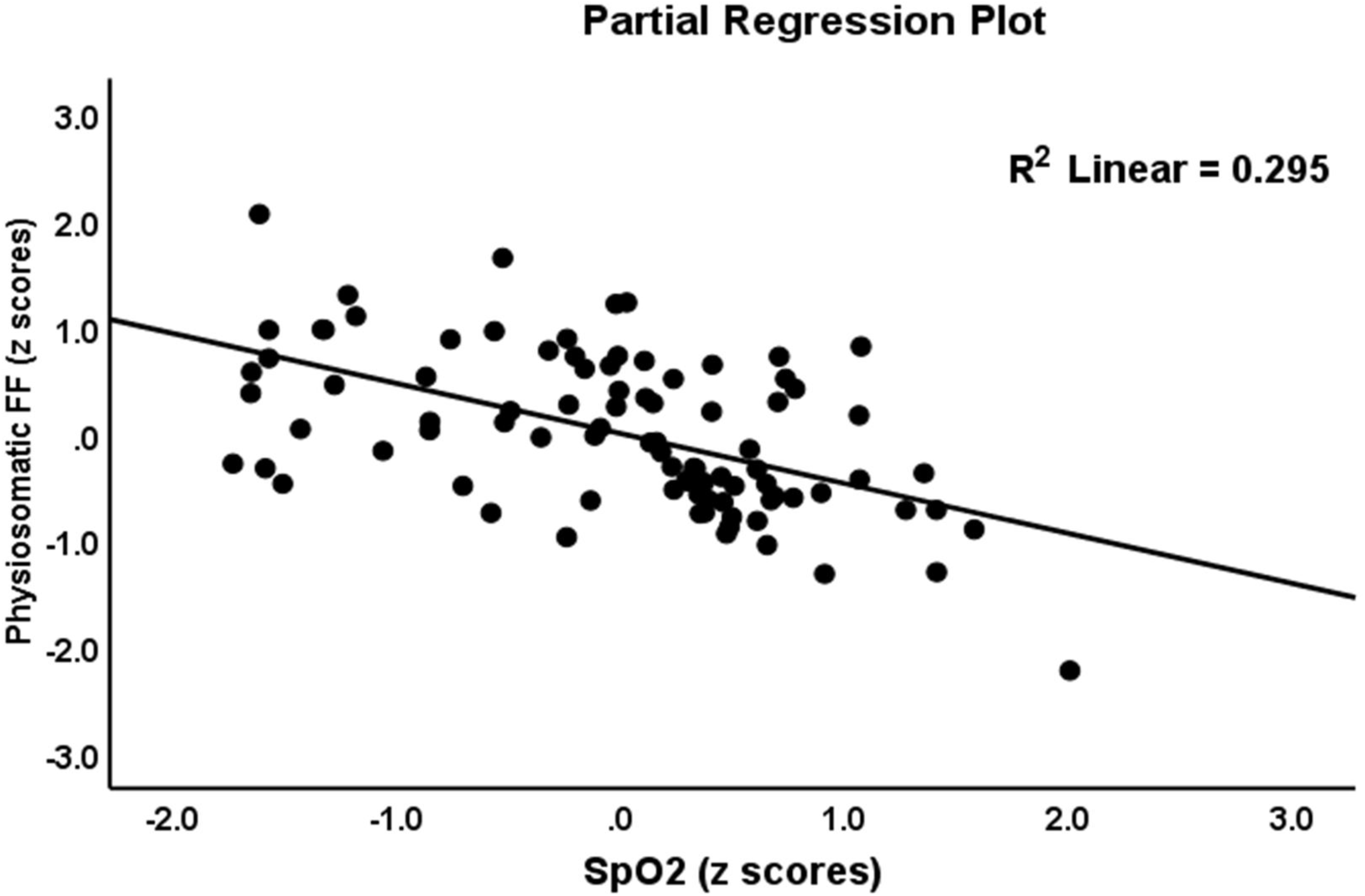
Partial regression of the pure physiosomatic component of the Fibromyalgia and Chronic Fatigue Rating Scale (FF) score on oxygen saturation percentage (SpO2)

**Table 5.** Results of multiple regression analysis with the Fibromyalgia and Chronic Fatigue Rating Scale (FF), total and subdomain, scores as dependent variables and chest CT scan anomalies (CCTAs), oxygen saturation percentage (SpO2), and biomarkers as explanatory variables.

| Dependent Variables | Explanatory Variables | $\beta$ | t | p | F model | df | p | R<sup>2</sup> |
| --- | --- | --- | --- | --- | --- | --- | --- | --- |
| #1. Total FF | Model |  |  |  | 51.64 | 3/86 | <0.001 | 0.643 |
|  | SpO2 | -0.478 | -6.32 | <0.001 |  |  |  |  |
|  | sRAGE | 0.271 | 2.99 | 0.004 |  |  |  |  |
|  | IL-6 | 0.223 | 2.66 | 0.009 |  |  |  |  |
| #2. Total FF | Model |  |  |  | 31.60 | 4/85 | <0.001 | 0.598 |
|  | sRAGE | 0.273 | 2.75 | 0.007 |  |  |  |  |
|  | Calcium | -0.341 | -3.41 | 0.001 |  |  |  |  |
|  | CRP | 0.189 | 2.17 | 0.033 |  |  |  |  |
|  | IL-6 | 0.192 | 2.12 | 0.037 |  |  |  |  |
| #3. Physiosomatic FF | Model |  |  |  | 45.933 | 3/86 | <0.001 | 0.616 |
|  | SpO2 | -0.470 | -6.00 | 0.000 |  |  |  |  |
|  | IL-6 | 0.246 | 2.83 | 0.006 |  |  |  |  |
|  | sRAGE | .237 | 2.52 | 0.014 |  |  |  |  |
| #4. Physiosomatic FF | Model |  |  |  | 37.00 | 3/86 | <0.001 | 0.563 |
|  | Calcium | -.399 | -4.63 | 0.000 |  |  |  |  |
|  | sRAGE | .263 | 2.60 | 0.011 |  |  |  |  |
|  | IL-6 | .240 | 2.58 | 0.011 |  |  |  |  |
| #5. Fatigue | Model |  |  |  | 37.33 | 2/87 | <0.001 | 0.462 |
|  | sRAGE | .451 | 4.91 | <0.001 |  |  |  |  |
|  | SpO2 | -.352 | -3.54 | 0.001 |  |  |  |  |
| #6. Fatigue | Model |  |  |  | 25.41 | 3/86 | <0.001 | 0.470 |
|  | sRAGE | .401 | 4.07 | <0.001 |  |  |  |  |
|  | CRP | .207 | 2.10 | 0.039 |  |  |  |  |
|  | Calcium | -.207 | -2.02 | 0.046 |  |  |  |  |

sRAGE: soluble advanced glycation end-product receptor, sACE2: soluble angiotensin converting enzyme 2, IL-6: interleukin (IL)-6, CRP: C-reactive protein.

#### Prediction of the HAM-A score

**Table 6** shows the results of multiple regressions of the HAM-A total and subdomain scores on the biomarker levels while allowing for the effects of demographic data. Regression #1 shows that 68.0% of the variance in the total HAM-A score could be explained by the regression on sRAGE and GGO (both positively), and SpO_2_ and calcium (both inversely). **Figure 3** shows the partial regression of the total HAM-A on sRAGE levels. The combination of sRAGE (positively) and calcium (negatively) explained 62.1% of the variance in the total HAM-A score (Regression #2). sRAGE and GGO explained 48.9% of the variance in the key HAM-A scores (Regression #3) and sRAGE and calcium explained 43.7% of the variance in the key HAM-A scores (regression #4). Regression #5 shows that sRAGE and GGO (both positively) and calcium (negatively) explained 41.9% of the variance in the physiosomatic HAM-A score. In regression #6 we found that 38.3% of the variance of the physiosomatic HAM-A scores could be explained by sRAGE (positively) and calcium (negatively). Finally, we also examined the regression of the sum of all physiosomatic symptoms on the biomarkers and found that sRAGE and GGO (positively) and SpO2 (inversely) explained 62.9% of its variance.

**Figure 3.**
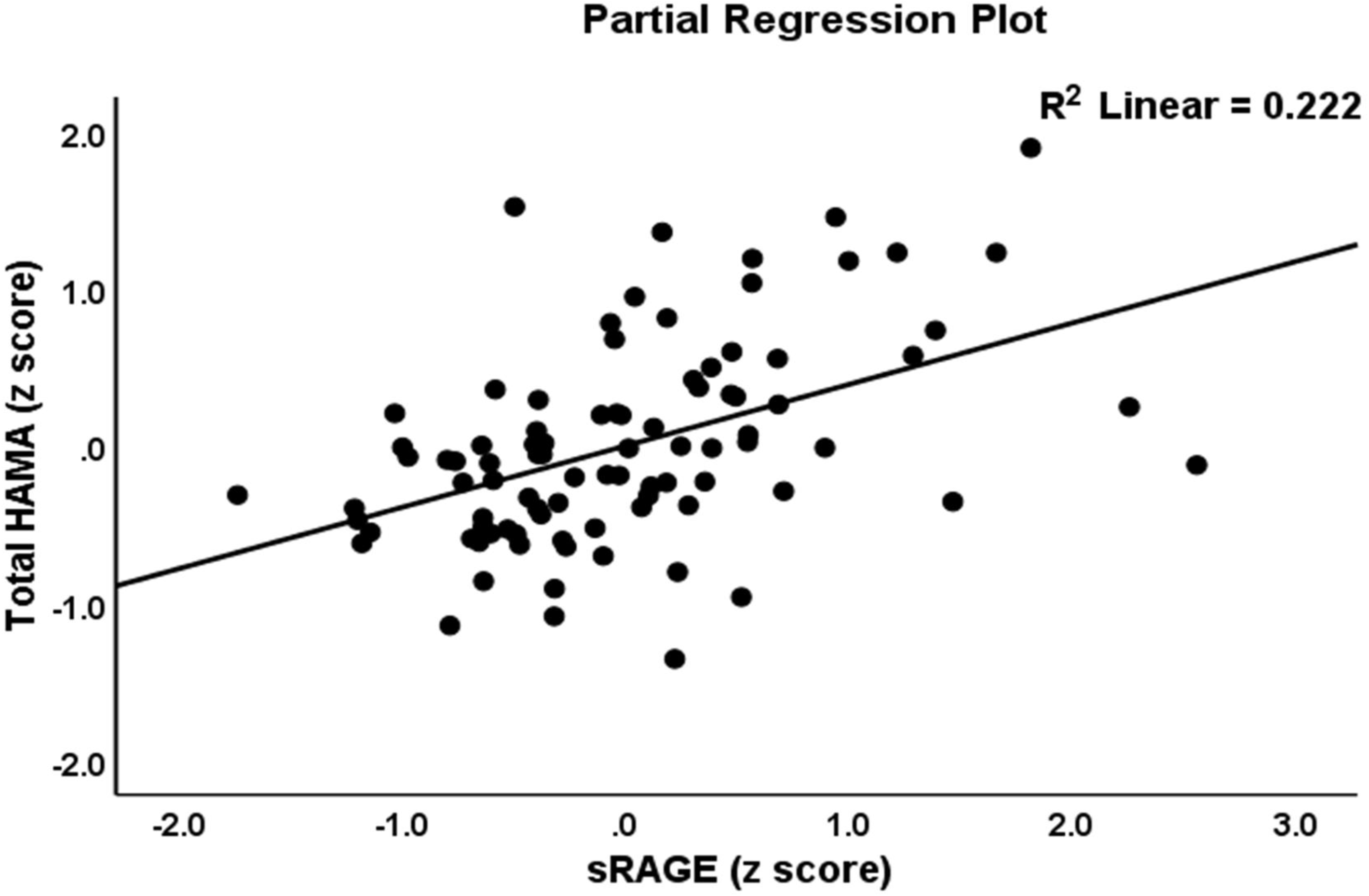
Partial regression of the total Hamilton Anxiety Rating Scale (HAMA) on levels of soluble receptor for advanced glycation products (sRAGE).

**Table 6.** Results of multiple regression analysis with the Hamilton Anxiety Rating Scale (HAM-A), total and subdomain, scores as dependent variables and chest CT scan anomalies (CCTAs), oxygen saturation percentage (SpO_2_), and biomarkers as explanatory variables.

| Dependent Variables | Explanatory Variables | $\beta$ | t | p | F model | df | p | R<sup>2</sup> |
| --- | --- | --- | --- | --- | --- | --- | --- | --- |
| #1. Total HAM-A | **Model** |  |  |  | 45.15 | 4/85 | <0.001 | 0.680 |
|  | sRAGE | 0.391 | 4.93 | <0.001 |  |  |  |  |
|  | SpO<sub>2</sub> | -0.220 | -2.35 | 0.021 |  |  |  |  |
|  | GGO | 0.192 | 2.23 | 0.029 |  |  |  |  |
|  | Calcium | -0.186 | -2.03 | 0.046 |  |  |  |  |
| #2. Total HAM-A | **Model** |  |  |  | 71.18 | 2/87 | <0.001 | 0.621 |
|  | sRAGE | 0.499 | 6.28 | <0.001 |  |  |  |  |
|  | Calcium | -0.392 | -4.93 | <0.001 |  |  |  |  |
| #3. Key HAM-A | **Model** |  |  |  | 41.61 | 2/87 | <0.001 | 0.489 |
|  | sRAGE | 0.433 | 4.66 | <0.001 |  |  |  |  |
|  | GGO | 0.356 | 3.82 | <0.001 |  |  |  |  |
| #4. Key HAM-A | **Model** |  |  |  | 33.77 | 2/87 | <0.001 | 0.437 |
|  | sRAGE | 0.511 | 5.28 | <0.001 |  |  |  |  |
|  | Calcium | -0.222 | -2.29 | 0.024 |  |  |  |  |
| #5. Physiosomatic HAM-A | **Model** |  |  |  | 20.67 | 3/86 | <0.001 | 0.419 |
|  | sRAGE | 0.301 | 2.85 | 0.006 |  |  |  |  |
|  | GGO | 0.249 | 2.29 | 0.024 |  |  |  |  |
|  | Calcium | -0.215 | -1.99 | 0.050 |  |  |  |  |
| #6. Physiosomatic HAM-A | **Model** |  |  |  | 27.06 | 2/87 | <0.001 | 0.383 |
|  | sRAGE | 0.388 | 3.83 | <0.001 |  |  |  |  |
|  | Calcium | -0.312 | -3.08 | 0.003 |  |  |  |  |
| #7. Sum all physiosomatic symptoms | **Model** |  |  |  | 48.69 | 3/86 | <0.001 | 0.629 |
|  | GGO | 0.301 | 3.32 | 0.001 |  |  |  |  |
|  | sRAGE | 0.335 | 4.06 | <0.001 |  |  |  |  |
|  | SpO<sub>2</sub> | -0.303 | -3.47 | 0.001 |  |  |  |  |

GGO: ground glass opacities, sRAGE: soluble advanced glycation end-product receptor, sACE2: soluble angiotensin converting enzyme 2, IL-6: interleukin (IL)-6, CRP: C-reactive protein.

### Results of PLS analyses

**Figure 4** shows a first PLS model conducted on 5.000 bootstrap samples. Melancholia, cognitive symptoms and insomnia could not be included in the same latent vector (due to low loadings) and, therefore, were entered as single indicators. One latent vector could be extracted from the key HDRS and HAM-A scores and the three psychosomatic domains as well (labeled physio-affective or PA-core). We were also able to combine all biomarkers into one latent vector (labeled as immune response), except sACE2 and magnesium which were entered as single indicators. We were able to extract one latent vector from CCTAs, crazy paving, consolidation, GGO, and other CCTAs, SpO2, and infection (a positive PCR test and IgM antibodies), labeled COVID-19 pneumonia. The construct reliabilities of the three latent vectors are good with AVE > 0.655, Cronbach α > 0.881, rho A > 0.873, and composite reliability > 0.904. The outer model loadings on the three latent vectors were > 0.721 at p<0.0001. The model fit was good with SRMR=0.050. CTA showed that the outer models were not mis-specified as reflective models. The construct cross-validated redundancies of the immune response (0.387) and PA-core (0.449) latent vectors were more than adequate. Full compositional invariance was obtained as indicated by the results of Prediction-Oriented Segmentation analysis, Measurement Invariance Assessment and Multi-Group Analysis. The Q_2_ Predict values of all construct indicators were positive which suggests that they outperform the most naïve benchmark. We found that 70.0% of the variance in the PA-core was explained by the regression on the immune response and pneumonia latent vectors. We found that the immune response explained 29.2% of the variance in melancholia and 9.7% of the variance in cognitive symptoms. Moreover, 28.7% of the variance in insomnia was explained by pneumonia and BMI (both positively associated). A large part the variance in the immune response (62.0%) was explained by the pneumonia latent vector. There were significant specific indirect effects of the latter on the PA-core (t=4.74, p<0.001), melancholia (t=7.50, p<0.001), cognitive symptoms (t=3.89, p<0.001), magnesium (t=4.50, p<0.001) and ACE2 (t=7.87, p<0.001), which were all mediated by the immune response latent vector.

**Figure 4.**
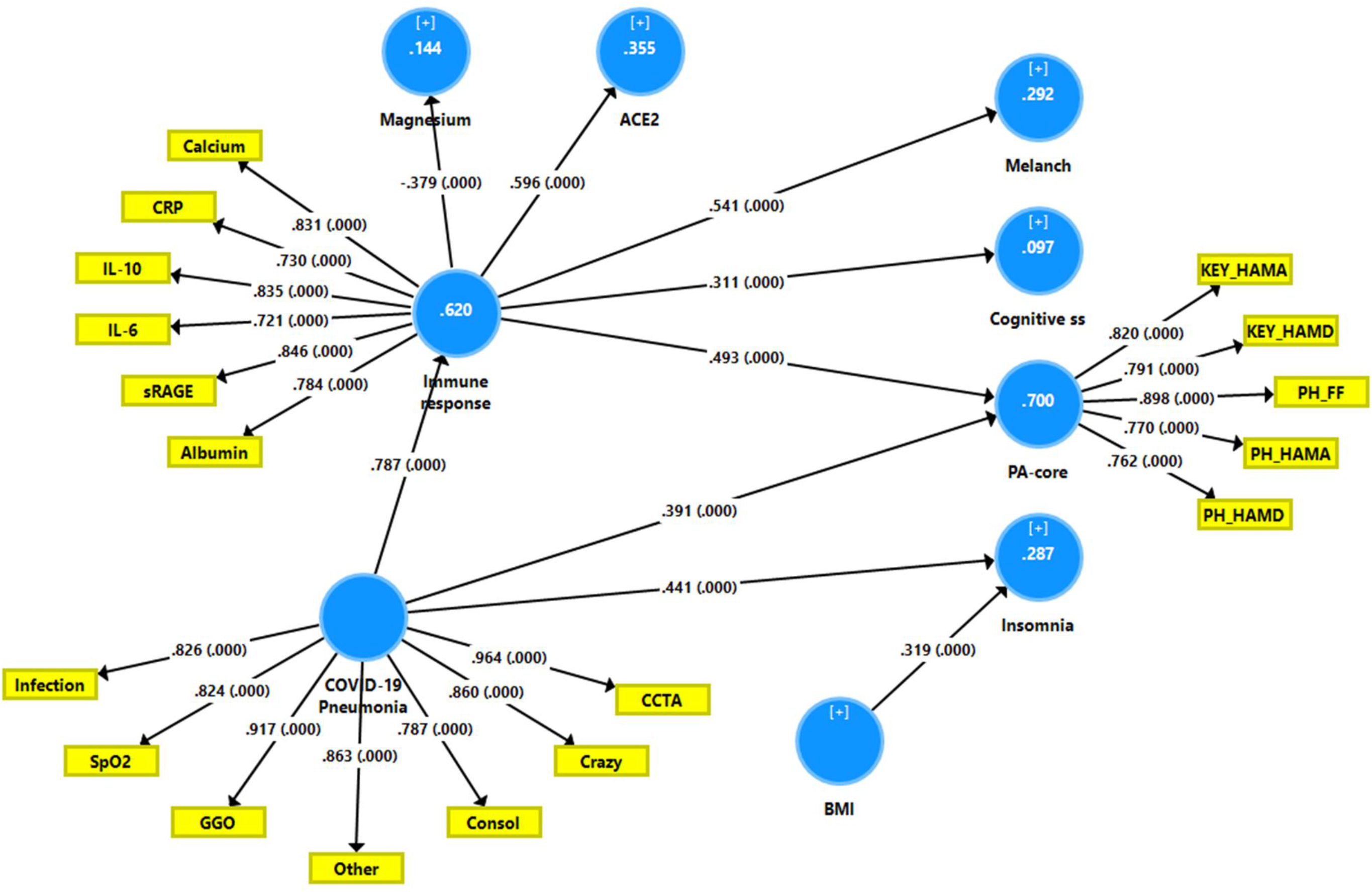
Results of Partial Least Squares (PLS)-SEM analysis with the PA-core (the physio-affective core), melancholic and cognitive symptoms and insomnia as output variables. The pathology of COVID-19 including chest CT scan anomalies (CCTAs), ground glass opacities (GGOs), crazy paving (crazy), consolidation (consol), other CCTAs (other) and lowered oxygen saturation (SpO2) as input variables. The immune response as indicated by a latent vector extracted from calcium, CRP (C-reactive protein), IL- 10 (interleukin-10), IL-6, sRAGEs (soluble receptor for advanced glycation end products) and albumin (partially) mediates the effects of COVID-19 on neuropsychiatric symptoms. Lowered magnesium and increased angiotensin converting enzyme 2 (ACE2) are spin- offs of the immune response. Figures in the circles indicate explained variance. Shown are path coefficients or latent vector loadings with accompanying p-values. KEY_HAMA: key anxiety symptoms of the Hamilton Anxiety Rating Scale. KEY_HAMD: key depressive symptoms of the Hamilton Depression Rating Scale. PH-HAMA/PH-HAMD: physiosomatic symptoms of HAMA/HAMD, respectively. PH_FF: physiosomatic symptoms of the Fibromyalgia and Chronic Fatigue Rating Scale (FF).

**Figure 5** shows a second PLS path analysis in which we have combined the immune response and pneumonia indicators into one latent vector named the infection-immune-inflammatory (III) core. The model fit (SRMR=0.051) was adequate and the construct reliability was adequate with AVE = 0.613, Cronbach α = 0.947, rho A > 0.954, and composite reliability > 0.953 and all III core loadings were all > 0.701 (except IL-6) at p<0.0001. This vector was not mis-specified as a reflective model. We found that this III core predicted the PA-core and other single indicator symptoms as well.

**Figure 5.**
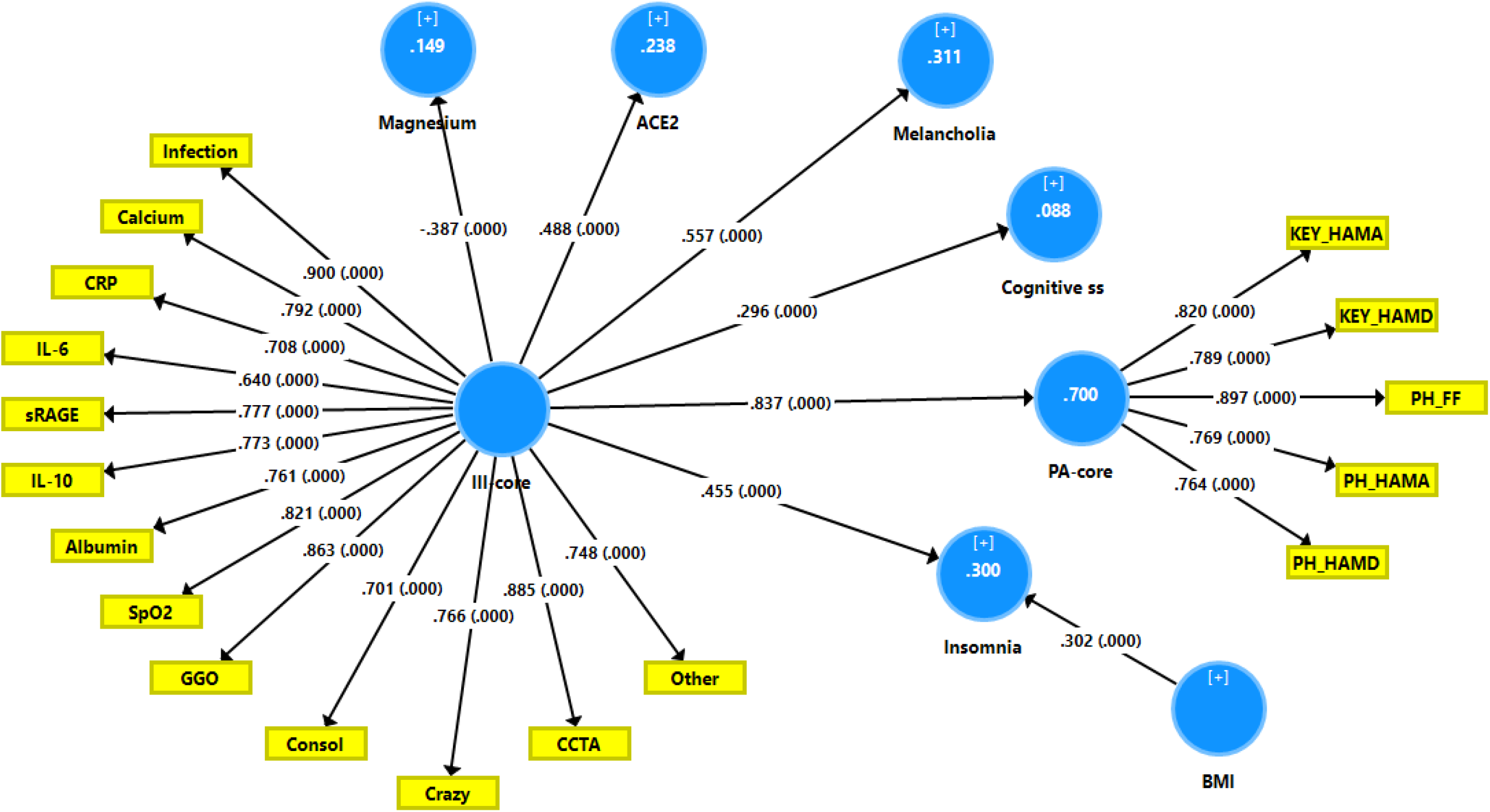
Results of Partial Least Squares (PLS)-SEM analysis with the PA-core (the physio-affective core), melancholic and cognitive symptoms and insomnia as output variables. Input variable is a latent vector named the infection-immune-inflammatory (III) core extracted from chest CT scan anomalies (CCTAs), ground glass opacities (GGOs), crazy paving (crazy), consolidation (consol), other CCTAs (other), lowered oxygen saturation (SpO2), calcium, CRP (C-reactive protein), IL-10 (interleukin-10), IL-6, sRAGEs (soluble receptor for advanced glycation end products) and albumin (partially). Lowered magnesium and increased angiotensin converting enzyme 2 (ACE2) are spin-offs of this III core but are not associated with neuro-psychiatric symptoms. Figures in the circles indicate explained variance. Shown are path coefficients or latent vector loadings with accompanying p-values. KEY_HAMA: key anxiety symptoms of the Hamilton Anxiety Rating Scale. KEY_HAMD: key depressive symptoms of the Hamilton Depression Rating Scale. PH-HAMA/PH-HAMD: physiosomatic symptoms of HAMA/HAMD, respectively. PH_FF: physiosomatic symptoms of the Fibromyalgia and Chronic Fatigue Rating Scale (FF).

### Discussion

#### COVID-19, affective and physiosomatic symptoms

A first major finding of this study is that COVID-19 is associated with increased levels of affective (including key depression and anxiety and melancholia) and physiosomatic FF symptoms as well as cognitive symptoms and insomnia. Because we have excluded patients with primary major depression, bipolar disorders and anxiety disorders, this association can best be described as depression, anxiety and physiosomatic (or ME/CFS-like) symptoms due to COVID-19. These findings extend the reports mentioned in our Introduction depicting the appearance of those symptoms in people with COVID-19. Depression was shown to be present in 8.3% to 48.3% of COVID-19 patients (Gao et al., 2020; Huang and Zhao, 2020; Ozamiz-Etxebarria et al., 2020). Other studies also reported increased levels of depression, distress, fear, sleep disorders, and suicidality in patients with COVID-19 (Luo et al., 2020; Qiu et al., 2020). Patients who are infected (or suspected of being infected) with COVID-19 can experience extreme emotional and behavioral reactions such as terror, boredom, isolation, anxiety, insomnia, or frustration (Shigemura et al., 2020). In COVID-19 patients, increased levels of fatigue are observed with a prevalence between 17.5% (Simani et al., 2021) and 53.6% (Qi et al., 2020). Chronic fatigue is also a key symptoms of the long (late) COVID-19 syndrome (Islam et al., 2020) (Décary et al., 2021).

Nevertheless, our study shows that in the acute phase of COVID-19, the appearance of affective (including key depressive and key anxiety symptoms) and physiosomatic FF symptoms is strongly interrelated and that this symptomatic response to COVID-19 is additionally characterized by the appearance of melancholia, cognitive symptoms, and insomnia. Furthermore, it appears that the key depressive, key anxiety and physiosomatic FF symptoms belong to a same underlying core and, therefore, that these symptoms are reflective manifestations of the same underlying phenomenon, which can best be described as the “physio-affective core”. It is interesting to note that such a core was also established in schizophrenia, major depression, ME/CFS, and somatoform disorder (Anderson and Maes, 2014; Kanchanatawan et al., 2019; Maes et al., 2021). On the other hand, melancholia, insomnia and cognitive symptoms do not belong to this common physio-affective core.

As described in the Introduction, the correlations between COVID-19 and affective and physiosomatic symptoms are often conceptualized as a consequence of psychological effects including the stigma that is associated with the infection, anxiety about possible adverse effects of the infection, the social isolation and reduction in social experiences, and the unemployment linked with quarantine or lockdown (Benke et al., 2020; Brooks et al., 2020; Kornilaki, 2021; Xiang et al., 2020). Nevertheless, as we will discuss in the following section, the physio-affective core, melancholic and cognitive symptoms, and insomnia are the consequence of infection, pneumonia, and the immune response in COVID-19.

#### Effects of pneumonia on affective and physiosomatic symptoms

The second major finding of this study is that the different CCTAs and lowered SpO2% were significantly associated with the appearance of the physio-affective core as well as with melancholia, cognitive symptoms and insomnia. Many (up to 70%) RT-PCR test positive COVID-19 patients show CCTAs (Adams et al., 2020), which indicate lung inflammation, bronchiolitis, pneumonia and lung fibrosis (Sadhukhan et al., 2020). In our study, the presence of CCTAs is strongly associated with lowered SpO2 indicating that pneumonia and lung lesions may cause decreased peripheral oxygen saturation, which is often decreased in COVID-19 patients and especially in those with more severe illness (Dai et al., 2020; Luks and Swenson, 2020). Silent hypoxia is one of the key symptoms of COVID-19, albeit it is not always the first symptom (Bouttell et al., 2020). In a recent study, the abnormalities in lung radiography such as presence of bilateral opacities, multifocal opacities, or any upper or middle zone opacity were associated with supplemental oxygen requirement (Ong et al., 2021).

There is some evidence that bronchitis and pneumonia are associated with major depressive symptoms (Adams et al., 2008; Seminog and Goldacre, 2013). In addition, patients with comorbid pneumonia and depression show a poorer treatment outcome as compared with patients without depression (Kao et al., 2014). On the other hand, depression is also a risk factor of hospitalization due to pneumonia (Davydow et al., 2014). According to the American Lung Association, low energy, fatigue, gastro-intestinal symptoms, neurocognitive impairments, and loss of appetite are typical symptoms of pneumonia (Niederman et al., 1993). Low blood oxygen levels or hypoxemia is also associated with depression and fatigue (Zhao et al., 2017).

#### The effects of pneumonia are partly mediated by immune activation

The third major finding of this study is that the effects of pneumonia and lowered SpO2 on the physio-affective core are partially mediated by the immune response and that pneumonia has also direct effects on this common core, suggesting that another process not mediated by immune activation may be involved. Furthermore, the effects of pneumonia on cognition and melancholia are completely mediated by immune activation. In the present study, the immune response was conceptualized as a common core underpinning the plasma levels of IL-6, IL-10, CRP, sRAGE (all increased), albumin and calcium (both decreased). Our results indicate that the acute phase and immune responses in COVID-19 are strongly associated with the physio-affective core. There is now evidence that affective disorders and ME/CFS are accompanied by an immune response (Bjørklund et al., 2020a; Gerwyn and Maes, 2017; Morris and Maes, 2013) and that the latter and its consequences may mechanistically explain symptoms of the physio-affective core and cognitive impairments (Kanchanatawan et al., 2019; Leonard and Maes, 2012; Morris and Maes, 2013). Interestingly, plasma levels of IL-6 and IL-10 in the acute phase of a virus infection have been shown to predict the progression of chronic fatigue (Russell et al., 2019).

In our study, lowered calcium is another component of the immune response latent vector in COVID-19 that is associated with the physio-affective core. Lowered calcium levels are frequently detected in COVID-19 with non-severe and severe illness (Di Filippo et al., 2021; Pal et al., 2020) and are often associated with severity of illness (Sun et al., 2020; Yang et al., 2021). Moreover, very low calcium values are associated with the severity of ARS (Sun et al., 2020) and the inflammatory response (Di Filippo et al., 2021; Di Filippo et al., 2020). Lowered calcium levels may be found during viral infections because albumin, which is lowered during the acute phase response, binds calcium and because viruses may utilize Ca^2+^ signals (Deng et al., 2012; Nieto-Torres et al., 2015; Zhou et al., 2009). Lowered calcium is frequently observed in patients with affective disorders and is associated with severity of depressive and physiosomatic symptoms (Al-Dujaili et al., 2019). Hypocalcemia is accompanied by a variety of physiosomatic symptoms including muscle tension, pain and cramps, cognitive impairments, and cardiovascular and respiratory symptoms (Bove-Fenderson and Mannstadt, 2018). Although magnesium is partly bound to albumin and is decreased in our COVID-19 patients, especially in those with extremely low SpO_2_ values, no associations with the physio-affective core or any other symptoms could be found after considering the role of the immune response. Nevertheless, magnesium deficiency may be accompanied by fatigue, lethargy, weakness, loss of appetite, numbness, muscle cramps, fibromyalgia-like symptoms, depression, and irritability (Ismail et al., 2018).

As described in the Introduction, binding of AGEs to the RAGEs on membranes initiates an immune-inflammatory response with elevated production of IL-6 and other cytokines (Macaione et al., 2007; Tobon-Velasco et al., 2014; Wang and Liu, 2016) and this response mediates not only inflammation, but also cell proliferation, migration, apoptosis, and microtubule stabilization (Xie et al., 2013) and this explains that the RAGE pathway is essential in COVID-19 progression (Yalcin Kehribar et al., 2021). The increased levels of sRAGEs in COVID-19 may be explained by proteolysis of the extracellular domain of RAGE (Sterenczak et al., 2009; Zhang et al., 2008), suggesting that increased sRAGE plasma levels in COVID-19 may reflect increased expression of membrane RAGEs. Interestingly, sRAGEs have anti-inflammatory properties by attenuating the binding to membrane RAGE (Oczypok et al., 2017; Sternberg et al., 2008; Yang et al., 2014). In major depression and bipolar disorder, sRAGE levels were significantly lower as compared with controls (Emanuele et al., 2011), suggesting that lowered levels could contribute to the immune response in mood disorders. As such, the increased sRAGE levels in our study are probably an indicant of the immune response in COVID- 19, rather than mediating the effects of pneumonia on the physio-affective core. Hypoxia may upregulate ACE2 gene expression and protein levels in lung and kidney which may contribute to the severity of COVID-19 (Shenoy et al., 2020). Nevertheless, the increased ACE2 levels established in our study were not associated with any affective or physiosomatic scores after considering the role of the immune response.

Importantly, our PLS analysis showed that one common “infection-immune-inflammatory core” underpins pneumonia-associated lung lesions, lowered SpO_2_ and immune activation, and that this core explains 70% of the variance in the physio-somatic core, and a relevant part of the variance in melancholia (31.1%), insomnia (30% when shared with BMI) and neurocognitive impairments (8.8%). As such, we may conclude that acute SARS-CoV-2 infection is often accompanied by lung lesions and lowered SpO2 which both are known to induce immune-inflammatory pathways (Sadhukhan et al., 2020) and that the increased incidence of neuro-psychiatric symptoms in COVID-19 should be attributed at least in part to the infection-immune-inflammatory core of COVID-19. Moreover, SARS-CoV-2 can infect the brain, causing neuroinflammation (Pan et al., 2020) and this is believed to be a another source of neuropsychiatric symptoms including chronic fatigue after recovery (Mandal et al., 2021).

#### Limitations

The results of the current study should be interpreted with regard to the limitations. First, this is a case-control study and, therefore, no firm causal relationships may be established. Second, it would have been even more interesting if we had assayed a set of neurotoxic immune biomarkers which are known to cause affective symptoms, including TNF-α and IL-1 signaling biomarkers, some chemokines, and oxidative stress biomarkers.

#### Conclusions

In COVID-19, one common core underpins lung lesions, lowered SpO2, and immune activation as indicated by increased plasma IL-6, IL-10, CRP, and sRAGE, and lowered albumin and calcium levels. This common “infection-immune-inflammatory core” explains a larger part of the variance in the physio-affective core, melancholic symptoms, insomnia, and neurocognitive complaints. Activated immune-inflammatory pathways mediate the effects of SARS-CoV-2 infection and pneumonia on the neuropsychiatric symptoms established in COVID-19.

## Data Availability

The dataset generated during and/or analyzed during the current study will be available from the corresponding author upon reasonable request and once the dataset has been fully exploited by the authors.

## Acknowledgements

The authors thank the staff of the CCU unit and the internal medicine clinic in Al-Sadr Teaching Hospital-Najaf City-Iraq for their help in the collection of samples. Also, we acknowledge the highly skilled work of the staff of Asia Laboratory in measuring the biomarkers.

## Declaration of interest

The authors have no financial conflict of interests.

## Funding

There was no specific funding for this specific study.

## Authorships

All authors contributed significantly to the paper and approved the final version.

## Notes

### Competing Interest Statement

The authors have declared no competing interest.

### Author Declarations

Institutional ethics board of the University of Kufa (617/2020).

## References

1. Adams, H.J., Kwee, T.C., Yakar, D., Hope, M.D., Kwee, R.M., 2020. Chest CT imaging signature of coronavirus disease 2019 infection: in pursuit of the scientific evidence. Chest 158, 1885–1895.

2. Adams, T.B., Wharton, C.M., Quilter, L., Hirsch, T., 2008. The association between mental health and acute infectious illness among a national sample of 18-to 24-year-old college students. J Amer Coll Health 56, 657–664.

3. Al-Dujaili, A.H., Al-Hakeim, H.K., Twayej, A.J., Maes, M., 2019. Total and ionized calcium and magnesium are significantly lowered in drug-naïve depressed patients: Effects of antidepressants and associations with immune activation. Metab Brain Dis 34, 1493–1503.

4. Al-Hakeim, H.K., Al-Jassas, H.K., Morris, G., Maes, M., 2021. Increased angiotensin-converting enzyme 2, sRAGE and immune activation, but lowered calcium and magnesium in COVID-19: association with chest CT abnormalities and lowered peripheral oxygen saturation. medRxiv, 2021.2003.2026.21254383.

5. Almulla, A.F., Al-Rawi, K.F., Maes, M., Al-Hakeim, H.K., 2021. In schizophrenia, immune-inflammatory pathways are strongly associated with depressive and anxiety symptoms, which are part of a latent trait which comprises neurocognitive impairments and schizophrenia symptoms. J Affect Disord 287, 316–326.

6. Anderson, G., Maes, M., 2014. Oxidative/nitrosative stress and immuno-inflammatory pathways in depression: treatment implications. Curr Pharm Des 20, 3812–3847.

7. Benjamini, Y., Hochberg, Y., 1995. Controlling the False Discovery Rate: A Practical and Powerful Approach to Multiple Testing. J R Stat Soc Series B Stat Methodol 57, 289–300.

8. Benke, C., Autenrieth, L.K., Asselmann, E., Pané-Farré, C.A., 2020. Lockdown, quarantine measures, and social distancing: Associations with depression, anxiety and distress at the beginning of the COVID-19 pandemic among adults from Germany. Psychiatry Res 293, 113462.

9. Bjørklund, G., Dadar, M., Pivina, L., Doşa, M.D., Semenova, Y., Maes, M., 2020a. Environmental, neuro-immune, and neuro-oxidative stress interactions in chronic fatigue syndrome. Molecular Neurobiology 57, 4598–4607.

10. Bjørklund, G., Dadar, M., Pivina, L., Doşa, M.D., Semenova, Y., Maes, M., 2020b. Environmental, neuro-immune, and neuro-oxidative stress interactions in chronic fatigue syndrome. Mol Neurobiol 57, 4598–4607.

11. Borges do Nascimento, I.J., Cacic, N., Abdulazeem, H.M., von Groote, T.C., Jayarajah, U., Weerasekara, I., Esfahani, M.A., Civile, V.T., Marusic, A., Jeroncic, A., Carvas Junior, N., Pericic, T.P., Zakarija-Grkovic, I., Meirelles Guimaraes, S.M., Luigi Bragazzi, N., Bjorklund, M., Sofi-Mahmudi, A., Altujjar, M., Tian, M., Arcani, D.M.C., O’Mathuna, D.P., Marcolino, M.S., 2020. Novel Coronavirus Infection (COVID-19) in Humans: A Scoping Review and Meta-Analysis. J Clin Med 9, 941.

12. Bouttell, J., Blane, D., Field, R., Heggie, R., Jani, B., Kelly, J., MacPherson, K., O’Donnell, K., Rana, D., Rattary, G., 2020. Assessment of COVID-19 in primary care: the identification of symptoms, signs, characteristics, comorbidities and clinical signs in adults which may indicate a higher risk of progression to severe disease.

13. Bove-Fenderson, E., Mannstadt, M., 2018. Hypocalcemic disorders. Best Pract Res Clin Endocrinol Metab 32, 639–656.

14. Brooks, S.K., Webster, R.K., Smith, L.E., Woodland, L., Wessely, S., Greenberg, N., Rubin, G.J., 2020. The psychological impact of quarantine and how to reduce it: rapid review of the evidence. Lancet 395, 912–920.

15. Cao, W., Fang, Z., Hou, G., Han, M., Xu, X., Dong, J., Zheng, J., 2020. The psychological impact of the COVID-19 epidemic on college students in China. Psychiatry Res 287, 112934.

16. Coronavirus-Resource-Center, 2021. Corona virus resource center. Johns Hopkins University & Medicine https://coronavirus.jhu.edu.

17. Dai, W.-C., Zhang, H.-W., Yu, J., Xu, H.-J., Chen, H., Luo, S.-P., Zhang, H., Liang, L.-H., Wu, X.-l., Lei, Y., 2020. CT imaging and differential diagnosis of COVID-19. Can Assoc Radiol J 71, 195–200.

18. Darif, D., Hammi, I., Kihel, A., El Idrissi Saik, I., Guessous, F., Akarid, K., 2021. The pro-inflammatory cytokines in COVID-19 pathogenesis: What goes wrong? Microb Pathog 153, 104799.

19. Davydow, D.S., Hough, C.L., Zivin, K., Langa, K.M., Katon, W.J., 2014. Depression and risk of hospitalization for pneumonia in a cohort study of older Americans. J Psychosom Res 77, 528–534.

20. Décary, S., Gaboury, I., Poirier, S., Garcia, C., Simpson, S., Bull, M., Brown, D., Daigle, F., 2021. Humility and Acceptance: Working Within Our Limits With Long COVID and Myalgic Encephalomyelitis/Chronic Fatigue Syndrome. J Orthop Sports Phys Ther 51, 197–200.

21. Deng, B., Zhang, S., Geng, Y., Zhang, Y., Wang, Y., Yao, W., Wen, Y., Cui, W., Zhou, Y., Gu, Q., 2012. Cytokine and chemokine levels in patients with severe fever with thrombocytopenia syndrome virus. PloS one 7, e41365.

22. Di Filippo, L., Formenti, A.M., Doga, M., Frara, S., Rovere-Querini, P., Bosi, E., Carlucci, M., Giustina, A., 2021. Hypocalcemia is a distinctive biochemical feature of hospitalized COVID-19 patients. Endocrine 71, 9–13.

23. Di Filippo, L., Formenti, A.M., Rovere-Querini, P., Carlucci, M., Conte, C., Ciceri, F., Zangrillo, A., Giustina, A., 2020. Hypocalcemia is highly prevalent and predicts hospitalization in patients with COVID-19. Endocrine 68, 475–478.

24. Emanuele, E., Martinelli, V., Carlin, M.V., Fugazza, E., Barale, F., Politi, P., 2011. Serum levels of soluble receptor for advanced glycation endproducts (sRAGE) in patients with different psychiatric disorders. Neurosci Lett 487, 99–102.

25. Fang Y, Z.H., Xie J, et al, 2020. Sensitivity of chest CT for COVID-19: comparison to RT-PCR. Radiology. Radiology 200432.

26. Franquet, T., 2011. Imaging of pulmonary viral pneumonia. Radiology 260, 18–39.

27. Gao, J., Zheng, P., Jia, Y., Chen, H., Mao, Y., Chen, S., Wang, Y., Fu, H., Dai, J., 2020. Mental health problems and social media exposure during COVID-19 outbreak. PLoS One 15, e0231924.

28. Gerwyn, M., Maes, M., 2017. Mechanisms Explaining Muscle Fatigue and Muscle Pain in Patients with Myalgic Encephalomyelitis/Chronic Fatigue Syndrome (ME/CFS): a Review of Recent Findings. Curr Rheumatol Rep 19, 1.

29. Gualano, M.R., Lo Moro, G., Voglino, G., Bert, F., Siliquini, R., 2020. Effects of Covid-19 lockdown on mental health and sleep disturbances in Italy. Int J Environ Res Public Health 17, 4779.

30. Hamilton, M., 1959. The assessment of anxiety states by rating. Br J Med Psychol 32, 50–55.

31. Hamilton, M., 1960. A rating scale for depression. J Neurol Neurosurg Psychiatry 23, 56–62.

32. Hansell, D.M., Bankier, A.A., MacMahon, H., McLoud, T.C., Muller, N.L., Remy, J., 2008. Fleischner Society: glossary of terms for thoracic imaging. Radiology 246, 697–722.

33. Huang, C., Wang, Y., Li, X., Ren, L., Zhao, J., Hu, Y., Zhang, L., Fan, G., Xu, J., Gu, X., Cheng, Z., Yu, T., Xia, J., Wei, Y., Wu, W., Xie, X., Yin, W., Li, H., Liu, M., Xiao, Y., Gao, H., Guo, L., Xie, J., Wang, G., Jiang, R., Gao, Z., Jin, Q., Wang, J., Cao, B., 2020. Clinical features of patients infected with 2019 novel coronavirus in Wuhan, China. Lancet 395, 497–506.

34. Huang, Y., Zhao, N., 2020. Generalized anxiety disorder, depressive symptoms and sleep quality during COVID-19 outbreak in China: a web-based cross-sectional survey. Psychiatry Res 288, 112954.

35. Hui, D.S.C., Zumla, A., 2019. Severe Acute Respiratory Syndrome: Historical, Epidemiologic, and Clinical Features. Infect Dis Clin North Am 33, 869–889.

36. Ismail, A.A.A., Ismail, Y., Ismail, A.A., 2018. Chronic magnesium deficiency and human disease; time for reappraisal? QJM 111, 759–763.

37. Kanchanatawan, B., Sriswasdi, S., Maes, M., 2019. Supervised machine learning to decipher the complex associations between neuro-immune biomarkers and quality of life in schizophrenia. Metab Brain Dis 34, 267–282.

38. Kanchanatawan, B., Thika, S., Sirivichayakul, S., Carvalho, A.F., Geffard, M., Maes, M., 2018. In Schizophrenia, Depression, Anxiety, and Physiosomatic Symptoms Are Strongly Related to Psychotic Symptoms and Excitation, Impairments in Episodic Memory, and Increased Production of Neurotoxic Tryptophan Catabolites: a Multivariate and Machine Learning Study. Neurotox Res 33, 641–655.

39. Kao, L.-T., Liu, S.-P., Lin, H.-C., Lee, H.-C., Tsai, M.-C., Chung, S.-D., 2014. Poor clinical outcomes among pneumonia patients with depressive disorder. PloS One 9, e116436.

40. Kornilaki, E.N., 2021. The psychological effect of COVID-19 quarantine on Greek young adults: Risk factors and the protective role of daily routine and altruism. Int J Psychol doi: 10.1002/ijop.12767.

41. Krishnan, A., Hamilton, J.P., Alqahtani, S.A., T, A.W., 2021. A narrative review of coronavirus disease 2019 (COVID-19): clinical, epidemiological characteristics, and systemic manifestations. Intern Emerg Med, 1-16.

42. Kwee, T.C., Kwee, R.M., 2020. Chest CT in COVID-19: What the Radiologist Needs to Know. RadioGraphics 40, 1848–1865.

43. Lambert, D.W., Yarski, M., Warner, F.J., Thornhill, P., Parkin, E.T., Smith, A.I., Hooper, N.M., Turner, A.J., 2005. Tumor necrosis factor-alpha convertase (ADAM17) mediates regulated ectodomain shedding of the severe-acute respiratory syndrome-coronavirus (SARS-CoV) receptor, angiotensin-converting enzyme-2 (ACE2). J Biol Chem 280, 30113–30119.

44. Leonard, B., Maes, M., 2012. Mechanistic explanations how cell-mediated immune activation, inflammation and oxidative and nitrosative stress pathways and their sequels and concomitants play a role in the pathophysiology of unipolar depression. Neurosci Biobehav Rev 36, 764–785.

45. Lindner, H.A., Velásquez, S.Y., Thiel, M., Kirschning, T., 2021. Lung Protection vs. Infection Resolution: Interleukin 10 Suspected of Double-Dealing in COVID-19. Front Immunol 12, 602130.

46. Liu, J., Li, S., Liu, J., Liang, B., Wang, X., Wang, H., Li, W., Tong, Q., Yi, J., Zhao, L., 2020a. Longitudinal characteristics of lymphocyte responses and cytokine profiles in the peripheral blood of SARS-CoV-2 infected patients. EBioMedicine 55, 102763.

47. Liu, Y., Yang, Y., Zhang, C., Huang, F., Wang, F., Yuan, J., Wang, Z., Li, J., Li, J., Feng, C., Zhang, Z., Wang, L., Peng, L., Chen, L., Qin, Y., Zhao, D., Tan, S., Yin, L., Xu, J., Zhou, C., Jiang, C., Liu, L., 2020b. Clinical and biochemical indexes from 2019-nCoV infected patients linked to viral loads and lung injury. Sci China Life Sci 63, 364–374.

48. Luks, A.M., Swenson, E.R., 2020. COVID-19 Lung Injury and High-Altitude Pulmonary Edema. A False Equation with Dangerous Implications. Ann Am Thorac Soc 17, 918–921.

49. Luo, Y., Kataoka, Y., Ostinelli, E.G., Cipriani, A., Furukawa, T.A., 2020. National prescription patterns of antidepressants in the treatment of adults with major depression in the US between 1996 and 2015: A population representative survey based analysis. Front Psychiat 11, 35.

50. Macaione, V., Aguennouz, M., Rodolico, C., Mazzeo, A., Patti, A., Cannistraci, E., Colantone, L., Di Giorgio, R.M., De Luca, G., Vita, G., 2007. RAGE-NF-kappaB pathway activation in response to oxidative stress in facioscapulohumeral muscular dystrophy. Acta Neurol Scand 115, 115–121.

51. Maes, M., 1993. Α review on the acute phase response in major depression. Rev Neurosci 4, 407–416.

52. Maes, M., Andres, L., Vojdani, A., Sirivichayakul, S., Barbosa, D.S., Kanchanatawan, B., 2021. In schizophrenia, chronic fatigue syndrome-and fibromyalgia-like symptoms are driven by breakdown of the paracellular pathway with increased zonulin and immune activation-associated neurotoxicity. medRxiv, doi: 10.1101/2021.1105.1109.21256897.

53. Maes, M., Bosmans, E., Suy, E., Vandervorst, C., De Jonckheere, C., Raus, J., 1990. Immune disturbances during major depression: upregulated expression of interleukin-2 receptors. Neuropsychobiol 24, 115–120.

54. Maes, M., Carvalho, A.F., 2018. The Compensatory Immune-Regulatory Reflex System (CIRS) in Depression and Bipolar Disorder. Mol Neurobiol 55, 8885–8903.

55. Maes, M., Scharpé, S., Meltzer, H.Y., Bosmans, E., Suy, E., Calabrese, J., Cosyns, P., 1993. Relationships between interleukin-6 activity, acute phase proteins, and function of the hypothalamic-pituitary-adrenal axis in severe depression. Psychiat Res 49, 11–27.

56. Maes, M., Twisk, F.N., 2010. Chronic fatigue syndrome: Harvey and Wessely’s (bio) psychosocial model versus a bio (psychosocial) model based on inflammatory and oxidative and nitrosative stress pathways. BMC medicine 8, 1–13.

57. Mandal, S., Barnett, J., Brill, S.E., Brown, J.S., Denneny, E.K., Hare, S.S., Heightman, M., Hillman, T.E., Jacob, J., Jarvis, H.C., 2021. ‘Long-COVID’: a cross-sectional study of persisting symptoms, biomarker and imaging abnormalities following hospitalisation for COVID-19. Thorax 76, 396–398.

58. Mehta, P., McAuley, D.F., Brown, M., Sanchez, E., Tattersall, R.S., Manson, J.J., 2020. COVID-19: consider cytokine storm syndromes and immunosuppression. Lancet 395, 1033–1034.

59. Montenegro, F., Unigarro, L., Paredes, G., Moya, T., Romero, A., Torres, L., Lopez, J.C., Gonzalez, F.E.J., Del Pozo, G., Lopez-Cortes, A., Diaz, A.M., Vasconez, E., Cevallos-Robalino, D., Lister, A., Ortiz-Prado, E., 2021. Acute respiratory distress syndrome (ARDS) caused by the novel coronavirus disease (COVID-19): a practical comprehensive literature review. Expert Rev Respir Med 15, 183–195.

60. Morris, G., Maes, M., 2013. Myalgic encephalomyelitis/chronic fatigue syndrome and encephalomyelitis disseminata/multiple sclerosis show remarkable levels of similarity in phenomenology and neuroimmune characteristics. BMC medicine 11, 1–23.

61. Niederman, M., Bass Jr, J., Campbell, G.D., Fein, A., Grossman, R., Mandell, L., Marrie, T., Sarosi, G., Torres, A., Yu, V., 1993. Guidelines for the initial management of adults with community-acquired pneumonia: diagnosis, assessment of severity, and initial antimicrobial therapy. American Thoracic Society. Medical Section of the American Lung Association. Am Rev Respir Dis 148, 1418–1426.

62. Nieto-Torres, J.L., Verdiá-Báguena, C., Jimenez-Guardeño, J.M., Regla-Nava, J.A., Castaño-Rodriguez, C., Fernandez-Delgado, R., Torres, J., Aguilella, V.M., Enjuanes, L., 2015. Severe acute respiratory syndrome coronavirus E protein transports calcium ions and activates the NLRP3 inflammasome. Virology 485, 330–339.

63. Oczypok, E.A., Perkins, T.N., Oury, T.D., 2017. All the "RAGE" in lung disease: The receptor for advanced glycation endproducts (RAGE) is a major mediator of pulmonary inflammatory responses. Paediatr Respir Rev 23, 40–49.

64. Ong, S.W.X., Hui, T.C.H., Lee, Y.S., Haja Mohideen, S.M., Young, B.E., Tan, C.H., Lye, D.C., 2021. High-risk chest radiographic features associated with COVID-19 disease severity. PLoS One 16, e0245518.

65. Oxley, T.J., Mocco, J., Majidi, S., Kellner, C.P., Shoirah, H., Singh, I.P., De Leacy, R.A., Shigematsu, T., Ladner, T.R., Yaeger, K.A., 2020. Large-vessel stroke as a presenting feature of Covid-19 in the young. N Engl J Med 382, e60.

66. Ozamiz-Etxebarria, N., Dosil-Santamaria, M., Picaza-Gorrochategui, M., Idoiaga-Mondragon, N., 2020. Niveles de estrés, ansiedad y depresión en la primera fase del brote del COVID-19 en una muestra recogida en el norte de España. Cad Saude Publica 36, 30.

67. Pal, R., Ram, S., Zohmangaihi, D., Biswas, I., Suri, V., Yaddanapudi, L.N., Malhotra, P., Soni, S.L., Puri, G.D., Bhalla, A., 2020. High prevalence of hypocalcemia in non-severe COVID-19 patients: a retrospective case-control study. Front Med 7, 590805.

68. Pan, F., Ye, T., Sun, P., Gui, S., Liang, B., Li, L., Zheng, D., Wang, J., Hesketh, R.L., Yang, L., 2020. Time course of lung changes on chest CT during recovery from 2019 novel coronavirus (COVID-19) pneumonia. Radiology 200370.

69. Pouya, M.A., Afshani, S.M., Maghsoudi, A.S., Hassani, S., Mirnia, K., 2020. Classification of the present pharmaceutical agents based on the possible effective mechanism on the COVID-19 infection. DARU, 1-20.

70. Qi, R., Chen, W., Liu, S., Thompson, P.M., Zhang, L.J., Xia, F., Cheng, F., Hong, A., Surento, W., Luo, S., Sun, Z.Y., Zhou, C.S., Li, L., Jiang, X., Lu, G.M., 2020. Psychological morbidities and fatigue in patients with confirmed COVID-19 during disease outbreak: prevalence and associated biopsychosocial risk factors. medRxiv doi: 10.1101/2020.05.08.20031666.

71. Qiu, J., Shen, B., Zhao, M., Wang, Z., Xie, B., Xu, Y., 2020. A nationwide survey of psychological distress among Chinese people in the COVID-19 epidemic: implications and policy recommendations. General psychiatry 33.

72. Ringle, C.M., Sarstedt, M., Straub, D.W., 2012. Editor’s comments: a critical look at the use of PLS-SEM in" MIS Quarterly". MIS quarterly 36, iii–xiv.

73. Russell, A., Hepgul, N., Nikkheslat, N., Borsini, A., Zajkowska, Z., Moll, N., Forton, D., Agarwal, K., Chalder, T., Mondelli, V., 2019. Persistent fatigue induced by interferon-alpha: a novel, inflammation-based, proxy model of chronic fatigue syndrome. Psychoneuroendocrinology 100, 276–285.

74. Sadhukhan, P., Ugurlu, M.T., Hoque, M.O., 2020. Effect of COVID-19 on Lungs: Focusing on Prospective Malignant Phenotypes. Cancers 12, 3822.

75. Seminog, O.O., Goldacre, M.J., 2013. Risk of pneumonia and pneumococcal disease in people with severe mental illness: English record linkage studies. Thorax 68, 171–176.

76. Shenoy, N., Luchtel, R., Gulani, P., 2020. Considerations for target oxygen saturation in COVID-19 patients: are we under-shooting? BMC Med 18, 260.

77. Shigemura, J., Ursano, R.J., Morganstein, J.C., Kurosawa, M., Benedek, D.M., 2020. Public responses to the novel 2019 coronavirus (2019-nCoV) in Japan: Mental health consequences and target populations. Psychiatry Clin Neurosci 74, 281–282.

78. Shmueli, G., Sarstedt, M., Hair, J.F., Cheah, J.-H., Ting, H., Vaithilingam, S., Ringle, C.M., 2019. Predictive model assessment in PLS-SEM: guidelines for using PLSpredict. Eur J Market 53 2322–2347.

79. Simani, L., Ramezani, M., Darazam, I.A., Sagharichi, M., Aalipour, M.A., Ghorbani, F., Pakdaman, H., 2021. Prevalence and correlates of chronic fatigue syndrome and post-traumatic stress disorder after the outbreak of the COVID-19. J Neurovirol 27, 154–159.

80. Sterenczak, K.A., Willenbrock, S., Barann, M., Klemke, M., Soller, J.T., Eberle, N., Nolte, I., Bullerdiek, J., Murua Escobar, H., 2009. Cloning, characterisation, and comparative quantitative expression analyses of receptor for advanced glycation end products (RAGE) transcript forms. Gene 434, 35–42.

81. Sternberg, D.I., Gowda, R., Mehra, D., Qu, W., Weinberg, A., Twaddell, W., Sarkar, J., Wallace, A., Hudson, B., D’Ovidio, F., Arcasoy, S., Ramasamy, R., D’Armiento, J., Schmidt, A.M., Sonett, J.R., 2008. Blockade of receptor for advanced glycation end product attenuates pulmonary reperfusion injury in mice. J Thorac Cardiovasc Surg 136, 1576–1585.

82. Sun, J.K., Zhang, W.H., Zou, L., Liu, Y., Li, J.J., Kan, X.H., Dai, L., Shi, Q.K., Yuan, S.T., Yu, W.K., Xu, H.Y., Gu, W., Qi, J.W., 2020. Serum calcium as a biomarker of clinical severity and prognosis in patients with coronavirus disease 2019. Aging (Albany NY) 12, 11287–11295.

83. Tanaka, T., Narazaki, M., Kishimoto, T., 2014. IL-6 in inflammation, immunity, and disease. Cold Spring Harb Perspect Biol 6, a016295.

84. Tobon-Velasco, J.C., Cuevas, E., Torres-Ramos, M.A., 2014. Receptor for AGEs (RAGE) as mediator of NF-kB pathway activation in neuroinflammation and oxidative stress. CNS Neurol Disord Drug Targets 13, 1615–1626.

85. Van Rheenen, T.E., Meyer, D., Neill, E., Phillipou, A., Tan, E.J., Toh, W.L., Rossell, S.L., 2020. Mental health status of individuals with a mood-disorder during the COVID-19 pandemic in Australia: Initial results from the COLLATE project. J Affect Disord 275, 69–77.

86. Vlachakis, D., Papakonstantinou, E., Mitsis, T., Pierouli, K., Diakou, I., Chrousos, G., Bacopoulou, F., 2020. Molecular mechanisms of the novel coronavirus SARS-CoV-2 and potential anti-COVID19 pharmacological targets since the outbreak of the pandemic. Food Chem Toxicol, 111805.

87. Wang, C., Pan, R., Wan, X., Tan, Y., Xu, L., Ho, C.S., Ho, R.C., 2020. Immediate psychological responses and associated factors during the initial stage of the 2019 coronavirus disease (COVID-19) epidemic among the general population in China. Int J Environ Res Public Health 17, 1729.

88. Wang, Y., Liu, L., 2016. The Membrane Protein of Severe Acute Respiratory Syndrome Coronavirus Functions as a Novel Cytosolic Pathogen-Associated Molecular Pattern To Promote Beta Interferon Induction via a Toll-Like-Receptor-Related TRAF3-Independent Mechanism. mBio 7, e01872–01815.

89. Xiang, Y.-T., Zhao, Y.-J., Liu, Z.-H., Li, X.-H., Zhao, N., Cheung, T., Ng, C.H., 2020. The COVID- 19 outbreak and psychiatric hospitals in China: managing challenges through mental health service reform. Int J Biol Sci 16, 1741.

90. Xiao, H., Zhang, Y., Kong, D., Li, S., Yang, N., 2020a. The effects of social support on sleep quality of medical staff treating patients with coronavirus disease 2019 (COVID-19) in January and February 2020 in China. Med Sci Mon Int Med J Exp Clin Res 26, e923549–923541.

91. Xiao, H., Zhang, Y., Kong, D., Li, S., Yang, N., 2020b. Social capital and sleep quality in individuals who self-isolated for 14 days during the coronavirus disease 2019 (COVID-19) outbreak in January 2020 in China. Med Sci Mon Int Med J Exp Clin Res 26, e923921–923921.

92. Xie, J., Méndez, J.D., Méndez-Valenzuela, V., Aguilar-Hernández, M.M., 2013. Cellular signalling of the receptor for advanced glycation end products (RAGE). Cell Signal 25, 2185–2197.

93. Yadav, R., Yadav, P., Kumar, S.S., Kumar, R., 2021. Assessment of Depression, Anxiety, and Sleep Disturbance in COVID-19 Patients at Tertiary Care Center of North India. J Neurosci Rural Pract 12, 316–322.

94. Yalcin Kehribar, D., Cihangiroglu, M., Sehmen, E., Avci, B., Capraz, A., Yildirim Bilgin, A., Gunaydin, C., Ozgen, M., 2021. The receptor for advanced glycation end product (RAGE) pathway in COVID-19. Biomarkers 26, 114–118.

95. Yang, C., Ma, X., Wu, J., Han, J., Zheng, Z., Duan, H., Liu, Q., Wu, C., Dong, Y., Dong, L., 2021. Low serum calcium and phosphorus and their clinical performance in detecting COVID-19 patients. J Med Virol 93, 1639–1651.

96. Yang, W.I., Lee, D., Lee, D.L., Hong, S.Y., Lee, S.H., Kang, S.M., Choi, D.H., Jang, Y., Kim, S.H., Park, S., 2014. Blocking the receptor for advanced glycation end product activation attenuates autoimmune myocarditis. Circ J 78, 1197–1205.

97. Zachrisson, O., Regland, B., Jahreskog, M., Kron, M., Gottfries, C.G., 2002. A rating scale for fibromyalgia and chronic fatigue syndrome (the FibroFatigue scale). J Psychosom Res 52, 501–509.

98. Zhang, H.-W., Yu, J., Xu, H.-J., Lei, Y., Pu, Z.-H., Dai, W.-C., Lin, F., Wang, Y.-L., Wu, X.-L., Liu, L.-H., 2020a. Corona virus international public health emergencies: implications for radiology management. Acad Radiol 27, 463–467.

99. Zhang, J., Lu, H., Zeng, H., Zhang, S., Du, Q., Jiang, T., Du, B., 2020b. The differential psychological distress of populations affected by the COVID-19 pandemic. Brain Behav Immun 87, 49–50.

100. Zhang, L., Bukulin, M., Kojro, E., Roth, A., Metz, V.V., Fahrenholz, F., Nawroth, P.P., Bierhaus, A., Postina, R., 2008. Receptor for advanced glycation end products is subjected to protein ectodomain shedding by metalloproteinases. J Biol Chem 283, 35507–35516.

101. Zhang, R., Wang, X., Ni, L., Di, X., Ma, B., Niu, S., Liu, C., Reiter, R.J., 2020c. COVID-19: Melatonin as a potential adjuvant treatment. Life Sci 250, 117583.

102. Zhao, F., Yang, J., Cui, R., 2017. Effect of Hypoxic Injury in Mood Disorder. Neural Plast 2017, 6986983.

103. Zhou, F., Yu, T., Du, R., Fan, G., Liu, Y., Liu, Z., Xiang, J., Wang, Y., Song, B., Gu, X., Guan, L., Wei, Y., Li, H., Wu, X., Xu, J., Tu, S., Zhang, Y., Chen, H., Cao, B., 2020. Clinical course and risk factors for mortality of adult inpatients with COVID-19 in Wuhan, China: a retrospective cohort study. Lancet 395, 1054–1062.

104. Zhou, Y., Frey, T.K., Yang, J.J., 2009. Viral calciomics: interplays between Ca2+ and virus. Cell Calcium 46, 1–17.

105. Zhu, N., Zhang, D., Wang, W., Li, X., Yang, B., Song, J., Zhao, X., Huang, B., Shi, W., Lu, R., 2020. A novel coronavirus from patients with pneumonia in China, 2019. N Engl J Med.

